# Food insecurity, caloric intake and nutritional status among children under 5 years old: a predictive modelling analysis of the MAL-ED multi-country cohort

**DOI:** 10.64898/2026.06.22.26355679

**Authors:** Francesco Checchi, Elaine Ferguson, Faisal Hamad, Yamna Ouchtar, Ruwan Ratnayake, Neha Singh, Hira Tanvir, Kevin van Zandvoort, Maysoon Dahab

## Abstract

**Background:** For children at risk of acute malnutrition, being able to predict and forecast dietary intakes and/or nutritional evolution would support decision-making, particularly in crisis settings where ground data collection is unfeasible or scant. We explored whether statistical models could offer accurate predictions of caloric intake or anthropometric (weight-for-height Z score, WHZ) changes, given intake, household food insecurity and other plausible predictors.

**Methods:** We reanalysed data from the Malnutrition and Enteric Disease (MAL-ED) multi-country (Bangladesh, Brazil, India, Nepal, Pakistan, Peru, South Africa, Tanzania) birth cohort (2009-2014), which consistently tracked household food insecurity experience, dietary intake, anthropometry, infectious disease symptoms, breastfeeding and other variables among children 9 to 35 months old. We quantified the performance on cross-validation of three models: (M1) change in WHZ as a function of household food insecurity; (M2) change in WHZ as a function of caloric intake; (M3) caloric intake as a function of household food insecurity. We compared random forests, lasso regressions, additive models and generalised boosted regressions. All models included age, sex, birth weight, urban versus rural residence, breastfeeding status and the longitudinal prevalence of diarrhoea, acute respiratory infection and fever as additional predictors.

**Results:** Altogether, M1, M2 and M3 leveraged 2957, 23,651 and 2013 longitudinal child observations, respectively. Both at country and individual level, there was low correlation among the key variables of interest. All three models featured low performance and moderate to extreme regression dilution, even when fitted to each country cohort separately.

**Discussion:** This secondary analysis based on data from a rigorous observational study suggests that statistical prediction of key variables along the causal pathway to childhood acute malnutrition may not be feasible. These negative findings may in part be explained by error in predictor measurement and the narrow range of both predictor and outcome values in the MAL-ED cohort, relative to the more extreme scenarios common to crisis settings. They also imply that mechanistic models requiring caloric intake as an input cannot rely on a statistical shortcut of this kind and must instead depend on empirical data or scenario assumptions.

## Background

In low-income settings, acute malnutrition underlies a large proportion of preventable child deaths [1]. Children affected by armed conflict, displacement, droughts and poly-crises experience food insecurity and a disproportionate burden of acute malnutrition [2]. Accordingly, to prevent child deaths decision-making on resource allocation and the design and monitoring of services in humanitarian responses should be predicated upon rapid but robust estimates of the prevalence and trends in both acute malnutrition and food insecurity [3], alongside other important determinants such as exposure to infections and infant and young child feeding practices (IYCF) [4].

In previous work [5], we showed that children’s weight trajectories and thus the population-level evolution of acute malnutrition prevalence can be projected using a mechanistic model. The necessary model include exposure to infectious disease (mainly diarrhoea and acute respiratory infections), breastfeeding status, treatment of severe (SAM) and/or moderate (MAM) acute malnutrition and, critically, caloric intake. These factors are captured by routine humanitarian surveys, assessments or monitoring data, except for caloric intake, which is notoriously difficult to measure, requiring lengthy, carefully implemented assessment methods such as 24-hour dietary recall [6, 7]. This may limit applicability of the above mechanistic model to theoretical scenario exploration, leaving cross-sectional ground surveys as the default solution for humanitarian monitoring, despite their cost and limited reach within insecure, hard-to-access populations [8, 9]. Critically, such surveys only capture nutritional deteriorations once they have happened: by contrast, forward-prediction through modelling would enable preventive actions.

It is generally understood that food insecurity, caloric intake and acute malnutrition are connected causally. This suggests that suitably parameterised statistical models might be able to issue predictions along this causal pathway. Such models, if sufficiently accurate, could predict the short-term evolution of malnutrition and provide inputs for mechanistic forecasts. However, prior findings on the association between food insecurity and acute malnutrition are mixed, and several systematic reviews have found weak or inconsistent relationships between the two [10–12] [refs]; We wished to test whether predictive modelling using a robust dataset of prospectively-collected data can offer better predictive accuracy. To do so, we reanalysed data from the Malnutrition and Enteric Disease Study (MAL-ED) multi-country cohort of children.

## Methods

### Data source

MAL-ED was conducted by a global partnership of researchers between 2009 and 2014, recruiting birth cohorts of children in eight countries (Bangladesh, Brazil, India, Nepal, Pakistan, Peru, South Africa, Tanzania), with longitudinal follow-up up to 59 months of age. The dataset has supported multiple analyses. Study methods have been extensively described [13], including those for infection- [14] and nutrition-related [15] measurements.

While none of the MAL-ED settings experienced bona fide crisis conditions (e.g. armed conflict), the study collected individual- and household-level data on food security, IYCF, infectious disease occurrence, caloric intake and anthropometry. These outcomes were measured with different frequencies over the child’s life course, as summarised in Table 1. Deviations from this intended schedule occurred due to individual child or country research team circumstances.

**Table 1.**
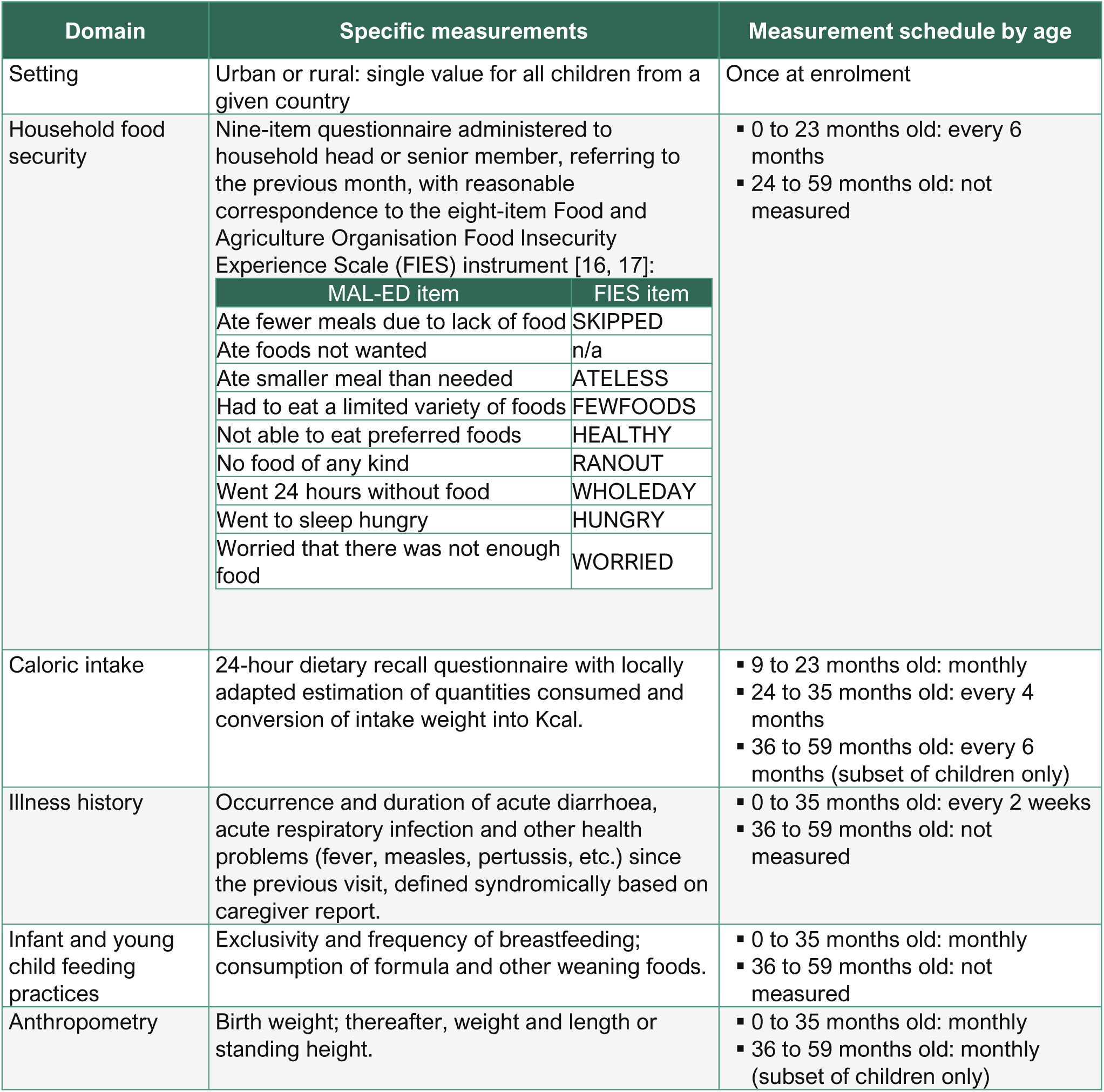
Intended schedule of measurements for specific MAL-ED cohort information domains relevant to this analysis.

### Pre-processing steps

We restructured the dataset around instances of anthropometric measurement, with other variables lined up around these instances to the extent available. We computed anthropometric indices (weight-for-height Z-score, WHZ; weight-for-age Z-score, WAZ) using the anthro package [18] per the WHO 2006 Growth Standards. Flags indicating possibly erroneous measurements were rare (64/74,623 for WHZ; 11/74,623 for WAZ) and we retained these as statistically plausible. We also computed the absolute daily mean change in WHZ since the previous measurement, excluding measurements taken > 40 days or < 20 days prior to maintain an approximately monthly data structure (i.e. while the change is expressed per day, it reflects inter-measurement periods of about one month).

We retained the MAL-ED classification of breastfeeding as exclusive, predominant, partial or none during the previous month. We computed the longitudinal prevalence of diarrhoea and ARI (proportion of observation days with illness symptoms) since the previous anthropometric measure, recoding prevalence as missing if data for <7 observation days were available. We also excluded caloric intake measurements and FIES questionnaires administered >7 days before or after the anthropometric measurement. We did not observe any clearly implausible caloric intake values (4/42,455 were > 4000 Kcal).

A method to compute globally comparable household-level probabilities and population-level prevalences of food insecurity from the eight-item FIES questionnaire has been developed [17]: the method recognises that cultural and other contextual factors shape households’ responses to the FIES items; it uses the single-parameter Rasch model to estimate the severity of each questionnaire item along a relative scale, and each household’s position within the same scale. We used its authors’ bespoke RM.weights package [19] to estimate household-level probabilities of experiencing any level of food insecurity, which we adopted as predictor for two of our models (M1 and M3 below). Notably, we recoded all item responses as binary (yes-no) and computed probabilities by country; while period stratification is recommended to avoid changes in response patterns over time, we wished to avoid data sparsity and also noted no marked seasonal or secular pattern in responses, though differences by country were obvious (Figure S1, Supplementary File). Rasch models did not converge for Brazil and Pakistan and, despite excluding outliers for specific food items (high ‘outfit’ statistic), model reliability (i.e. the modelled country-specific scale’s discriminatory power) remained low for Nepal (< 0.70). Accordingly, only FIES observations from Bangladesh, India, Peru, South Africa and Tanzania were retained. Estimated item severities for each country were standardised by linear transformation to those of the global reference scale developed by Cafiero et al. [20], using as anchor points questionnaire items from each country model with a severity within ±0.35 of that of the global scale. Lastly, probabilities of the household being moderately or severely food-insecure were computed based on the household’s raw FIES score (number of items with a ‘yes’ response), as shown by Viviani [21].

### Statistical analysis

We tested the predictive accuracy of three models:

- **M1:** Food insecurity as predictor of anthropometric (WHZ) change;
- **M2:** Caloric intake as predictor of anthropometric (WHZ) change;
- **M3:** Food insecurity as predictor of caloric intake.

For each model, we included other categorical (breastfeeding status; sex; urban or rural setting) and continuous (longitudinal prevalence of diarrhoea and ARI; birth weight; age) predictors. While the MAL-ED cohort contains a wealth of other variables (e.g. detailed dietary intake; aetiology of diarrhoea), we wished to restrict predictors to those likely to be available via surveys and routine data collection in real-life contexts where the above models might be useful. Availability of data for each model is summarised in Table 2, and Figure S2 reports variable availability by age.

**Table 2.** Data attrition and availability of data for each model.

| Pre-processing step | Number of child observations<br>(% of starting number) | Model |
| --- | --- | --- |
| Initial set of anthropometric observations with a preceding value, enabling calculation of anthropometric change outcomes since the previous measurement (differences in weight, WHZ and WAZ) | 64,860 (100.0%) |  |
| Remove observations with missing country, urban/rural residence and/or birth weight | 57,455 (88.6%) |  |
| Remove observations with missing breastfeeding exclusivity | 45,916 (70.8%) |  |
| Remove observations with missing longitudinal prevalence of disease (<7 days with disease observations since the previous anthropometric measurement) | 43,042 (66.4%) |  |
| Remove observations for which the preceding anthropometric measurement occurred more than 40 days or fewer than 20 days earlier (set A) | 42,717 (65.9%) |  |
| Remove observations without a caloric intake measure within $\pm 7$ days (set B) | 23,651 (36.5%) | M2 |
| Remove observations without a probability of the household being food insecure as assessed by a FIES questionnaire within $\pm 7$ days†: | | |
| from set A | 2,957 (4.6%) | M1 |
| from set B | 2,013 (3.1%) | M3 |

† All observations from Brazil, Nepal and Pakistan were excluded.

For each model we tested the following prediction methods: (i) a random forest (using the ranger package [22]), grown with 500 trees and a maximum three variables to split data on at every node (values up to five of this parameter were tested but were less accurate); (ii) a lasso regularisation regression to optimise inclusion of predictors (using glmnet [23]), with the shrinkage parameter determined through exploratory cross-validation [24]; (iii) a generalised additive mixed model [25] (using mgcv [26]) with child ID as a random effect and p-spline smoothing terms for the main predictor of interest and child’s age (due to computational intractability, random effects consisting of child ID nested into country were not included, and the random effect was removed altogether for M2); (iv) a generalised boosted regression model (using gbm [27]) with optimal values for the shrinkage parameter, tree complexity and number of trees determined through exploratory cross-validation. All four methods assumed a Gaussian distribution of the outcome; caloric intake was square-root transformed to approach normality (Figure S3).

The out-of-sample performance of each method was evaluated using 100-fold cross-validation, in which a given fraction (‘fold’) of children (1% for 100 folds) was held out of the training data, with this fraction distributed across the country cohorts proportionally to their size, and all observations from a given child attributed to the training or validation set; the model was trained on remaining data and predictive accuracy on the hold-out fold was tracked using root mean square error (RMSE) and absolute bias. After cycling through all possible folds, the model’s mean RMSE and bias were evaluated. We also plotted predictions versus observations in individual folds to graphically assess performance. Sensitivity analyses were carried out by fitting each model to individual country datasets.

### Ethics and data availability

Source datasets cannot be shared, and must instead be requested from the MAL-ED investigators (see https://clinepidb.org/ce/app/workspace/analyses/DS_5c41b87221/new/details). However, code with which to replicate the analysis in R software [28] on these source datasets is available at https://github.com/francescochecchi/amber_maled_analysis. This secondary analysis was approved by the London School of Hygiene and Tropical Medicine Ethics Committee (ref. 32330).

## Results

Table 3 summarises key characteristics of each model dataset. For most countries data for approximately 200-250 individual children were available. There were remarkable country differences in the main variables of interest: notably, at this aggregate level there was little correlation among food insecurity prevalence, caloric intake and WHZ, with Bangladeshi children in particular featuring relatively low probability of food insecurity despite a lower mean WHZ and median caloric intake than Peruvian and South African children where the probability of food insecurity was higher and mean WHZ above 0. Generally, malnutrition was higher in the M2 than M1 dataset, possibly due to a higher median age in the former. Bi-variate correlations between predictor-outcome sets are shown in Figure S4 (M1), Figure S10 (M2) and Figure S16 (M3).

**Table 3.** Characteristics of children included in each model, by country and overall.

| Characteristic | Bangladesh | Brazil | India | Nepal | Pakistan | Peru | S. Africa | Tanzania | Total |
| --- | --- | --- | --- | --- | --- | --- | --- | --- | --- |
| **M1: Food insecurity → anthropometric change** |  |  |  |  |  |  |  |  |  |
| **N observations (children)** | 740<br>(229) | n/a | 620<br>(229) | n/a | n/a | 812<br>(259) | 497<br>(228) | 288<br>(119) | **2957**<br>**(1064)** |
| **Female children (%)** | 370<br>(50.0%) | n/a | 324<br>(52.3%) | n/a | n/a | 371<br>(45.7%) | 239<br>(48.1%) | 141<br>(49.0%) | **1445**<br>**(48.9%)** |
| **Median age in months (IQR§)** | 12.1 (6.1 to 18.1) | n/a | 12.1 (6.0 to 18.1) | n/a | n/a | 12.1 (6.1 to 18.1) | 17.9 (12.0 to 24.0) | 12.0 (6.0 to 18.0) | **12.1 (6.1 to 18.1)** |
| **Percent food insecure†** | 1.4% | n/a | 6.8% | n/a | n/a | 10.4% | 12.1% | 1.9% | **6.9%** |
| **Mean WHZ (IQR)** | -0.59 (-1.23 to 0.07) | n/a | -0.84 (-1.50 to -0.24) | n/a | n/a | 0.46 (-0.23 to 1.16) | 0.38 (-0.35 to 1.09) | 0.34 (-0.36 to 1.06) | **-0.14 (-0.89 to 0.71)** |
| **N acutely malnourished (%)‡** | 51 (6.9%) | n/a | 73 (11.8%) | n/a | n/a | 10 (1.2%) | 11 (2.2%) | 2 (0.7%) | **147 (5.0%)** |
| **M2: Caloric intake → anthropometric change** |  |  |  |  |  |  |  |  |  |
| **N observations (children)** | 3435<br>(228) | 1848<br>(197) | 2977<br>(226) | 4880<br>(224) | 438<br>(69) | 5151<br>(243) | 3386<br>(249) | 1536<br>(117) | **23,651**<br>**(1553)** |
| **Female children (%)** | 1661<br>(48.4%) | 868<br>(47.0%) | 1601<br>(53.8%) | 2237<br>(45.8%) | 240<br>(54.8%) | 2334<br>(45.3%) | 1721<br>(50.8%) | 806<br>(52.5%) | **11,468**<br>**(48.5%)** |
| **Median age in months (IQR)** | 18.0 (13.0 to 22.9) | 17.1 (13.1 to 22.1) | 17.9 (13.0 to 22.1) | 20.0 (14.0 to 26.0) | 27.9 (23.3 to 32.0) | 21.0 (14.1 to 28.1) | 18.9 (14.0 to 23.0) | 19.9 (14.1 to 25.0) | **19.0 (14.0 to 24.1)** |
| **Median Kcal intake (IQR)** | 352 (208 to 556) | 1023 (750 to 1277) | 769 (496 to 1053) | 507 (282 to 804) | 999 (730 to 1244) | 858 (540 to 1195) | 907 (626 to 1195) | 1048 (803 to 1301) | **732 (416 to 1078)** |
| **Mean WHZ (IQR)** | -0.72 (-1.33 to -0.06) | 0.65 (-0.20 to 1.66) | -0.93 (-1.55 to -0.33) | -0.25 (-0.80 to 0.32) | -1.10 (-1.61 to -0.51) | 0.30 (-0.34 to 0.91) | 0.36 (-0.34 to 1.04) | 0.24 (-0.45 to 0.90) | **-0.13 (-0.86 to 0.63)** |
| **N acutely malnourished (%)‡** | 295 (8.6%) | 42 (2.3%) | 366 (12.3%) | 97 (2.0%) | 72 (16.4%) | 76 (1.5%) | 70 (2.1%) | 14 (0.9%) | **1032**<br>**(4.4%)** |
| **M3: Food in security → caloric intake** |  |  |  |  |  |  |  |  |  |
| **N observations (children)** | 540<br>(222) | n/a | 352<br>(200) | n/a | n/a | 570<br>(232) | 372<br>(205) | 179<br>(103) | **2013**<br>**(962)** |
| **Female children (%)** | 272<br>(50.4%) | n/a | 194<br>(55.1%) | n/a | n/a | 258<br>(45.3%) | 184<br>(49.5%) | 89<br>(49.7%) | **997**<br>**(49.5%)** |
| **Median age in months (IQR)** | 18.0 (12.0 to 24.0) | n/a | 18.0 (12.0 to 24.0) | n/a | n/a | 18.0 (12.1 to 24.0) | 18.0 (12.1 to 24.0) | 17.9 (12.0 to 18.1) | **18.0 (12.1 to 24.0)** |
| **Percent food insecure†** | 1.5% | n/a | 5.1% | n/a | n/a | 9.9% | 10.3% | 2.0% | **6.2%** |
| **Median Kcal intake (IQR)** | 335 (204 to 515) | n/a | 764 (508 to 1063) | n/a | n/a | 736 (473 to 1032) | 914 (673 to 1175) | 1018 (802 to 1252) | **689 (394 to 1013)** |

§ Inter-quartile range. † Mean probability of the household experiencing any level of food insecurity, across all observations. ‡ Observations with WHZ < -2.

All models and prediction methods had low performance (Table 4), with evidence of considerable bias, error and regression dilution (biassing of the predicted-versus-observed slope towards zero [29]). Figure 1, Figure 2, Figure 3 show prediction-versus-observation plots on cross-validation for M1, M2 and M3 respectively. Low performance was observed even when excluding Bangladesh observations (given the country cohort’s notably low mean caloric intake relative to other settings), and when fitting models to each individual country’s dataset (Figures S9, S15, S21): these sensitivity analyses were done using random forests only, given this method yielded the best combination of bias and error across the three models. Of the three models, M3 (food insecurity → caloric intake) featured the most promising performance, provided regression dilution could be corrected (see Discussion).

**Figure 1.**
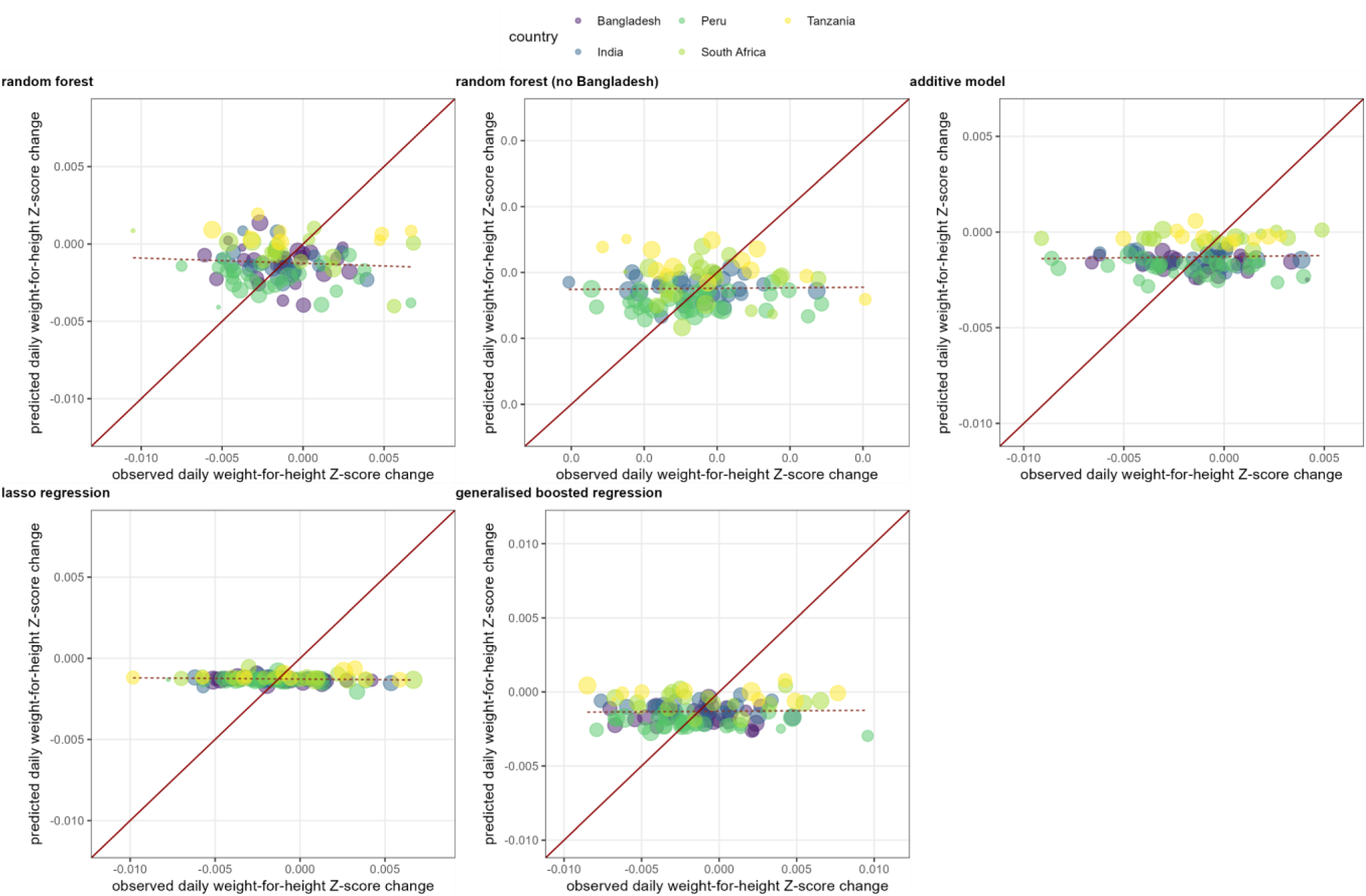
Mean predictions versus observations, by cross-validation fold and prediction method, for M1. Each dot represents a cross-validation fold, sized according to its relative share of the total dataset. The diagonal line indicates optimal prediction, while the dotted line indicates the slope of the predictions as a function of observations, based on a least-squares regression weighted by fold size.

**Figure 2.**
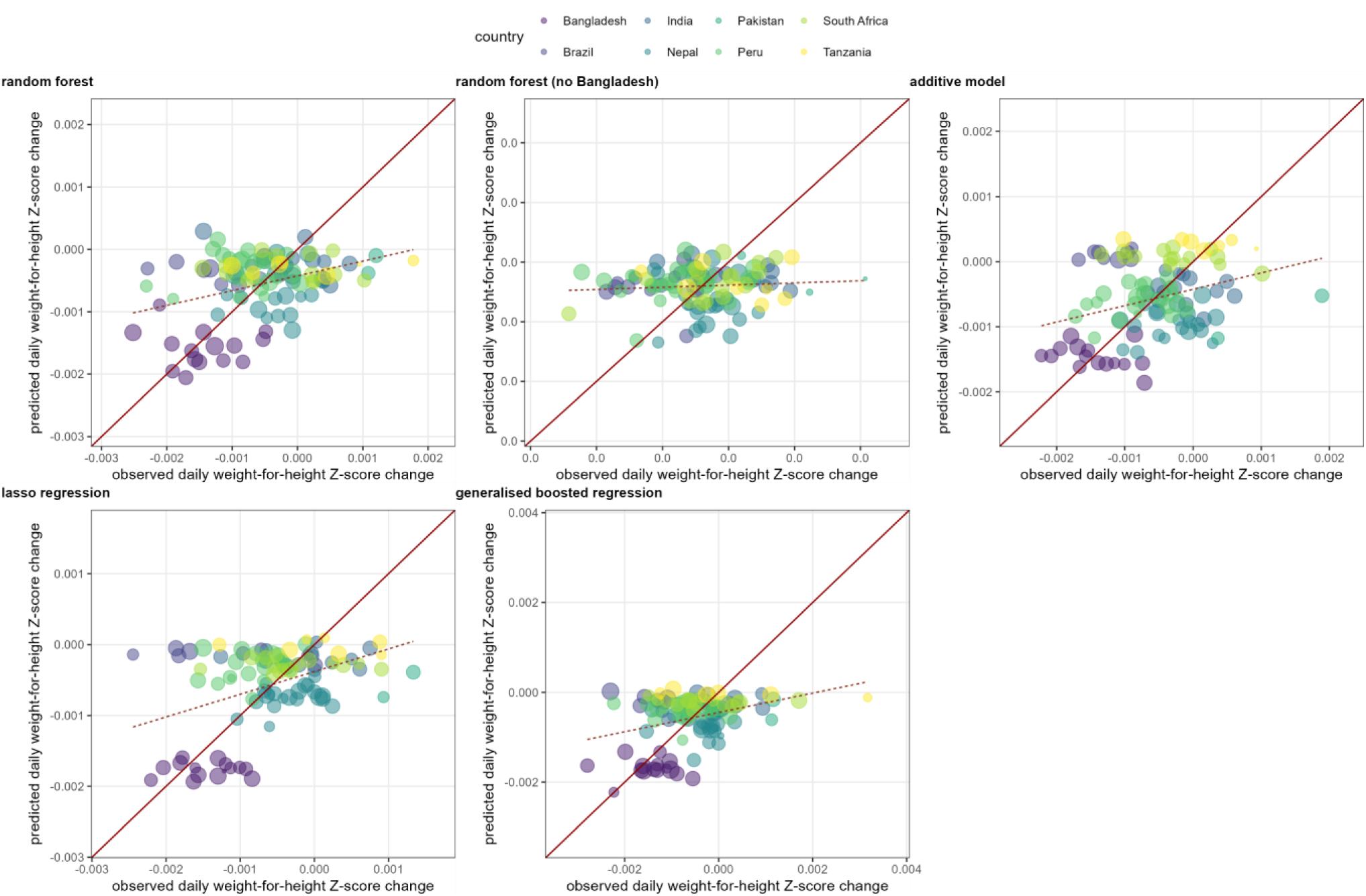
Mean predictions versus observations, by cross-validation fold and prediction method, for M2. Each dot represents a cross-validation fold, sized according to its relative share of the total dataset. The diagonal line indicates optimal prediction, while the dotted line indicates the slope of the predictions as a function of observations, based on a least-squares regression weighted by fold size.

**Figure 3.**
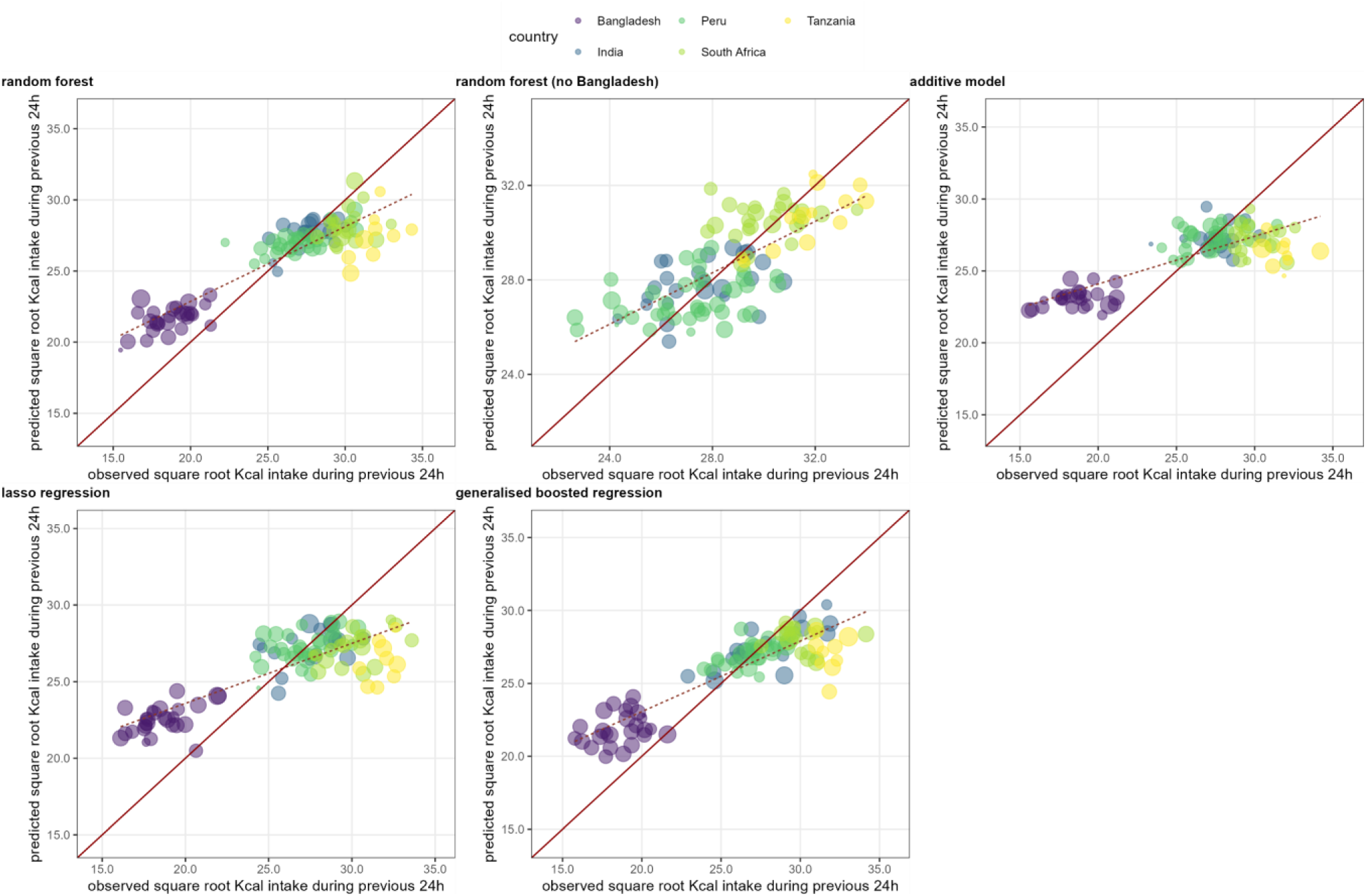
Mean predictions versus observations, by cross-validation fold and prediction method, for M3. Each dot represents a cross-validation fold, sized according to its relative share of the total dataset. The diagonal line indicates optimal prediction, while the dotted line indicates the slope of the predictions as a function of observations, based on a least-squares regression weighted by fold size.

**Table 4.**
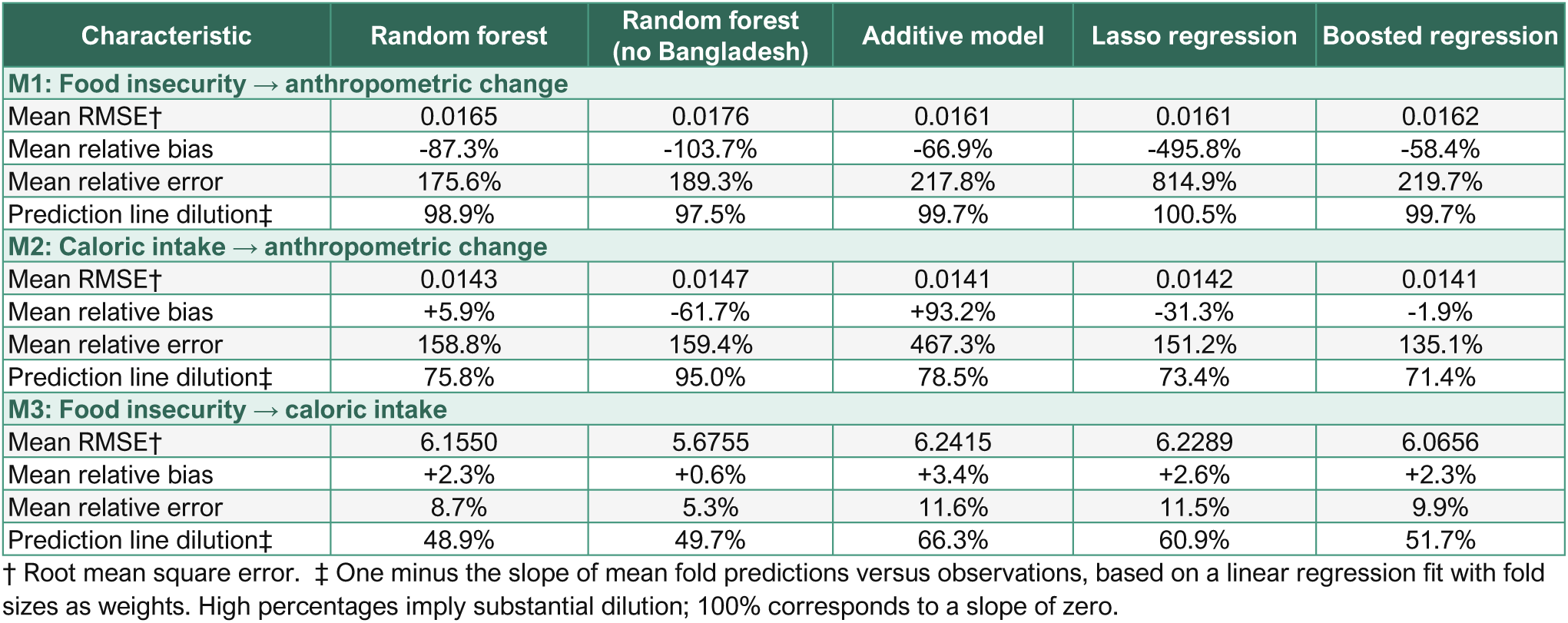
Summary performance of each tested prediction method on cross-validation, by model. All models were tested on 100 cross-validation folds and included the following other predictors: urban vs. rural, birth weight, age, sex, breastfeeding status, longitudinal prevalence of ARI, longitudinal prevalence of diarrhoea, longitudinal prevalence of fever.

| Characteristic | Random forest | Random forest (no Bangladesh) | Additive model | Lasso regression | Boosted regression |
| --- | --- | --- | --- | --- | --- |
| **M1: Food insecurity → anthropometric change** |  |  |  |  |  |
| Mean RMSE† | 0.0165 | 0.0176 | 0.0161 | 0.0161 | 0.0162 |
| Mean relative bias | -87.3% | -103.7% | -66.9% | -495.8% | -58.4% |
| Mean relative error | 175.6% | 189.3% | 217.8% | 814.9% | 219.7% |
| Prediction line dilution‡ | 98.9% | 97.5% | 99.7% | 100.5% | 99.7% |
| **M2: Caloric intake → anthropometric change** |  |  |  |  |  |
| Mean RMSE† | 0.0143 | 0.0147 | 0.0141 | 0.0142 | 0.0141 |
| Mean relative bias | +5.9% | -61.7% | +93.2% | -31.3% | -1.9% |
| Mean relative error | 158.8% | 159.4% | 467.3% | 151.2% | 135.1% |
| Prediction line dilution‡ | 75.8% | 95.0% | 78.5% | 73.4% | 71.4% |
| **M3: Food insecurity → caloric intake** |  |  |  |  |  |
| Mean RMSE† | 6.1550 | 5.6755 | 6.2415 | 6.2289 | 6.0656 |
| Mean relative bias | +2.3% | +0.6% | +3.4% | +2.6% | +2.3% |
| Mean relative error | 8.7% | 5.3% | 11.6% | 11.5% | 9.9% |
| Prediction line dilution‡ | 48.9% | 49.7% | 66.3% | 60.9% | 51.7% |

† Root mean square error. ‡ One minus the slope of mean fold predictions versus observations, based on a linear regression fit with fold sizes as weights. High percentages imply substantial dilution; 100% corresponds to a slope of zero.

Random forest and generalised boosted regression methods provide relative measures of predictor importance or influence, respectively: these are useful to quantify how much of the outcome’s variability can be explained by a predictor of interest. Despite slight disagreement, both methods suggested that food insecurity was less important than birth weight and age for M1 (food insecurity → WHZ change; Figures S6-7), and, for M3 (food insecurity → caloric intake), much less important than breastfeeding status (by far the most important predictor), age and birth weight (Figures S18-19). However, caloric intake was the most important predictor for M2 (caloric intake → WHZ change; Figures S12-13).

We also visualised the extent to which each key predictor influenced the outcome by outputting model predictions at different hypothetical levels of the key predictor, while holding all other predictor values at the levels in the dataset; specifically, we varied the probability of food insecurity from 0% to 100%, and caloric intake by multiplying the values in the data by factors of 0.5 to 1.5. As expected, scenarios with higher food insecurity featured lower or negative predicted WHZ change (M1, Figure S8); similarly, as intake increased, so did predicted WHZ change (M2, Figure S14).

However, predicted intake was insensitive to varying scenarios of food insecurity (M3, Figure S20), as expected given the latter’s relatively low importance as a predictor. Note that these partial dependence plots may look similar to but should not be confused with dose-response plots arising from a causal association model, which would have been fitted by adjusting for plausible confounders in a search for parsimony, and which has a purpose distinct from that of a predictive model.

## Discussion

This re-analysis of the MAL-ED dataset suggests that accurate statistical prediction of anthropometric change or caloric intake in children as a function of household food insecurity, caloric intake itself and other plausible predictors may be difficult or prohibitive, even when working with data from a carefully executed growth cohort. None of the models evaluated featured a performance high enough to support their practical use, e.g. as a substitute for ground data collection.

Several studies have reported high performance of statistical models predicting or forecasting food insecurity, with armed conflict and environmental variables noted as highly predictive (e.g. see [30–32]). We previously explored statistical prediction of acute malnutrition burden in crisis settings, with training data at the individual level but a range of predictors measured at the population level, including drought conditions, food purchasing power, epidemic occurrence and insecurity. Models had poor performance in both South Sudan and Somalia [33], but showed some promise in drought-prone areas of Kenya, with less bias than observed in this analysis [34]. An entirely population-level analysis at administrative level 2 across 36 Sub-Saharan African countries reported reasonable forecasting performance for acute malnutrition prevalence based on environmental and conflict intensity variables, but the outcome was not available with sufficient frequency to enable prediction of the evolution of malnutrition over short time intervals; instead, models were put forward as supporting broad early warning [35].

The poor predictive value of food insecurity for acute malnutrition in this study is in line with a pilot cross-sectional sample ahead of MAL-ED cohort implementation [36] and several systematic reviews [37–39]: we speculate that this is due to measurement error and to the complex and probably non-linear interplay of food insecurity, coping mechanisms and other factors driving malnutrition. Though we did not conduct a systematic search, we found few studies, predictive or causal, considering the link between caloric intake and acute malnutrition, or with intake as the outcome. MAL-ED investigators found that caloric intake from complementary food was associated with growth rates at 24 months of age [40], as borne out by our dependence plots (Figures S12-13). In Niger, dietary quality but not Kcal amount was associated with WHZ < -2 [41]. It is likely that the disappointing performance of M2 in our study (caloric intake → anthropometric change) reflects in part other (e.g. infectious disease aetiology and severity) differences across children and countries in the cohort. In particular, the strong influence of breastfeeding status suggests that the model largely picks up differences in successful weaning transition within our cohort of young children.

### Limitations

While the MAL-ED study undertook painstaking data collection based on standardised protocols, predictor and outcome data included in this analysis are likely to suffer from varying levels of measurement error. In particular, infectious disease occurrence relied on caregiver report and syndromic case definitions; anthropometric measurements are error-prone, especially length in young children; the 24-hour recall method for measuring caloric intake is complicated to administer, relies on local and approximate measurements of food quantities, and is generally known to yield inaccuracy; the FIES questionnaire also relies on accurate responses by households and, despite the method replicated here to standardise food insecurity variables across different countries, may feature unaccounted-for biases, e.g. due to perceptions of desirable responses by households. Moreover, its conversion to household-level probabilities of food insecurity rests on several assumptions, and the probabilities themselves, while treated deterministically in our model, feature uncertainty. Taken together, it is plausible that the food insecurity predictor analysed here is only partially reflective of objective food insecurity conditions experienced by households, and subject to large measurement error. Moreover, there was a timeframe mismatch between food insecurity and WHZ change on the one hand (both measured with reference to the previous month) and caloric intake on the other: the latter measure reflected only a 24h period around the end of the same month, likely featuring high random variation relative to a month-long measure due to various factors such as illness, care circumstances, metabolism, etc. This alone may explain poor model performance to a considerable extent.

The sum total of predictor error would typically be expected to cause regression dilution. The prediction-versus-observation graphs do indeed suggest extensive dilution for M1 and M2 and some for M3. While methods for correcting for prediction slope attenuation have been proposed, these require carrying out repeat measurements at least in a sub-sample of study participants, so as to estimate measurement variability [42].

While the MAL-ED cohort was longitudinal, two predictors/outcomes of interest (caloric intake, food insecurity) were assessed only at certain time points, reducing the effective sample sizes of M1 and M3 and the range of ages at which these were evaluated. Though MAL-ED encompassed a range of settings across LMICs, as demonstrated by the variability in the main predictors and outcomes, it is possible that the most severe end of the spectrum of food insecurity and malnutrition was not reflected in the composition of the study cohorts, depriving this analysis of observations akin those often found in crisis-affected populations, where food insecurity in particular may reach such severe levels that households exhaust all available coping mechanisms [43], resulting in a far more pronounced effect on caloric intake and anthropometry: otherwise put, this analysis may cover that portion of the full spectrum of food insecurity for which effects on childhood malnutrition may be largely mitigated through adaptation and/or caloric sacrifice by adults, but not levels at which children experience prolonged starvation. Indeed, acute malnutrition prevalence was relatively high in South Asia but very low elsewhere (Table 3). MAL-ED also did not collect middle-upper arm circumference data, which are influential for humanitarian assessments, or data on interventions such as management of infectious disease episodes or malnutrition itself.

### Conclusions

This secondary analysis indicates that statistical prediction of the evolution of nutritional status among children, or of their caloric intake, may not be a viable approach in settings where such predictions would support decision-making. It is possible that more accurate models could be developed based on datasets representative of settings with more extreme variability in food insecurity and caloric intakes, but this would require ad-hoc and rigorous primary, prospective data collection to generate a suitable model training and validation dataset composed of a wide spectrum of predictors related to food security, coping, disease incidence, and treatment measured longitudinally at closer time points. A key, possibly intractable challenge remains how to measure predictors with sufficient accuracy to reduce regression dilution and thus underestimation of the outcome: if this challenge affected MAL-ED measurements, it would likely be even more problematic in crisis settings, unless a large-scale study to measure predictors and outcomes were accompanied by repeat measurements of predictors particularly subject to error: this would enable correction of regression dilution. Mechanistic models or expert elicitation approaches, perhaps combined with limited ground measurement, may hold more promise as an approach for anticipating the evolution of acute malnutrition in crisis settings to inform decisions. Notably, however, such models require caloric intake as an input [5], meaning that without a statistical shortcut of the kind tested here, they must rely either on resource-intensive dietary recall data, which are often infeasible in crisis settings, or on untested scenario assumptions. Such approaches would also require rigorous validation in multiple settings.

## Data Availability

Source datasets cannot be shared, but are available upon reasonable request from the MAL-ED investigators. Code with which to replicate the analysis in R software on these source datasets is available.

https://clinepidb.org/ce/app/workspace/analyses/DS_5c41b87221/new/details

https://github.com/francescochecchi/amber_maled_analysis

## Acknowledgements

This study was funded by UK aid from the UK government; however the views expressed do not necessarily reflect the UK government’s official policies. We are grateful to Zeina Jamaluddine for advising on the FIES preprocessing steps.

