## Supplementary File for "Food insecurity, caloric intake and nutritional status among children under 5 years old: a predictive modelling analysis of the MAL-ED multi-country cohort"

### Table of figures

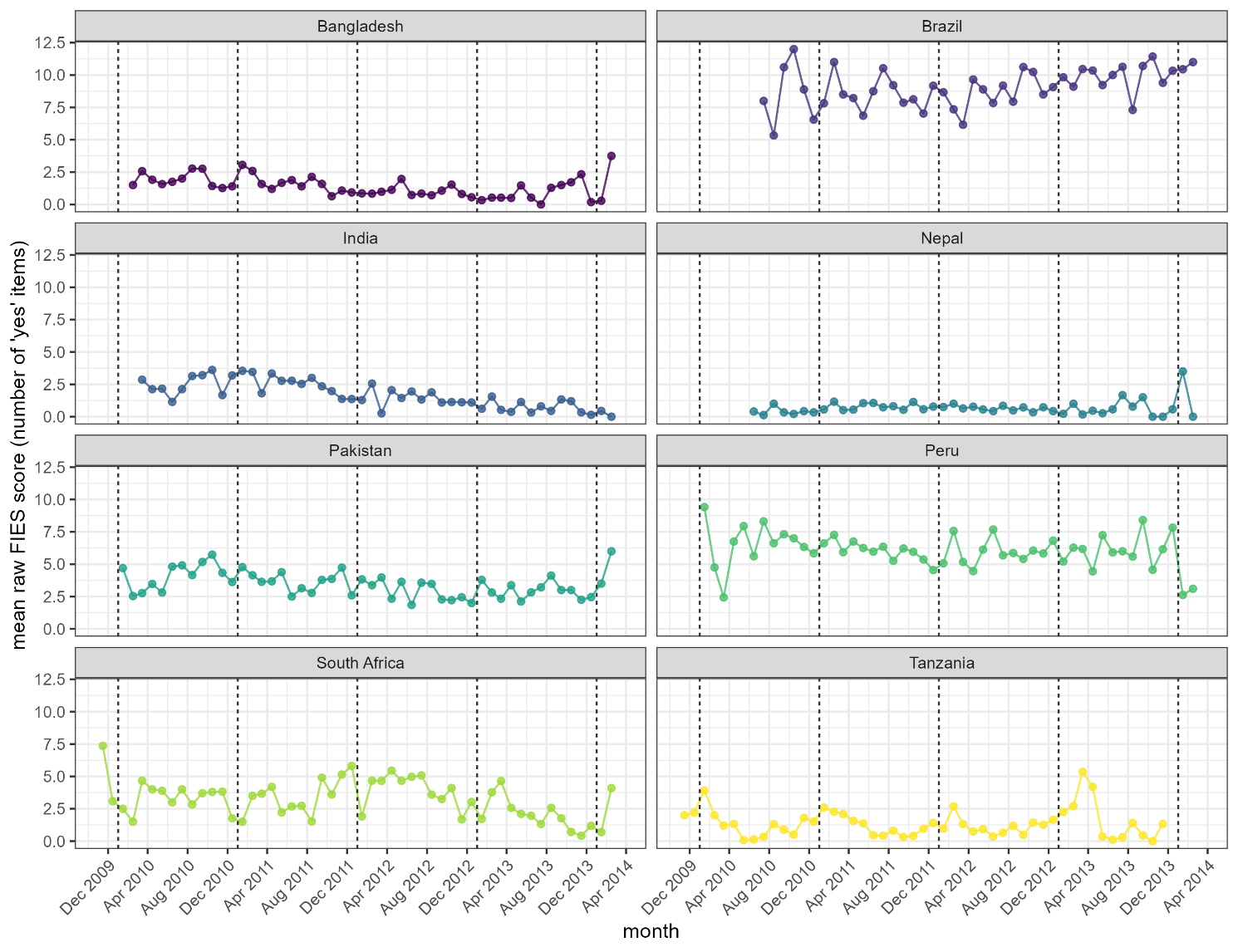

Figure S1. Mean crude FIES score (sum of ‘yes’ answers to the eight-item questionnaire), by country and week.

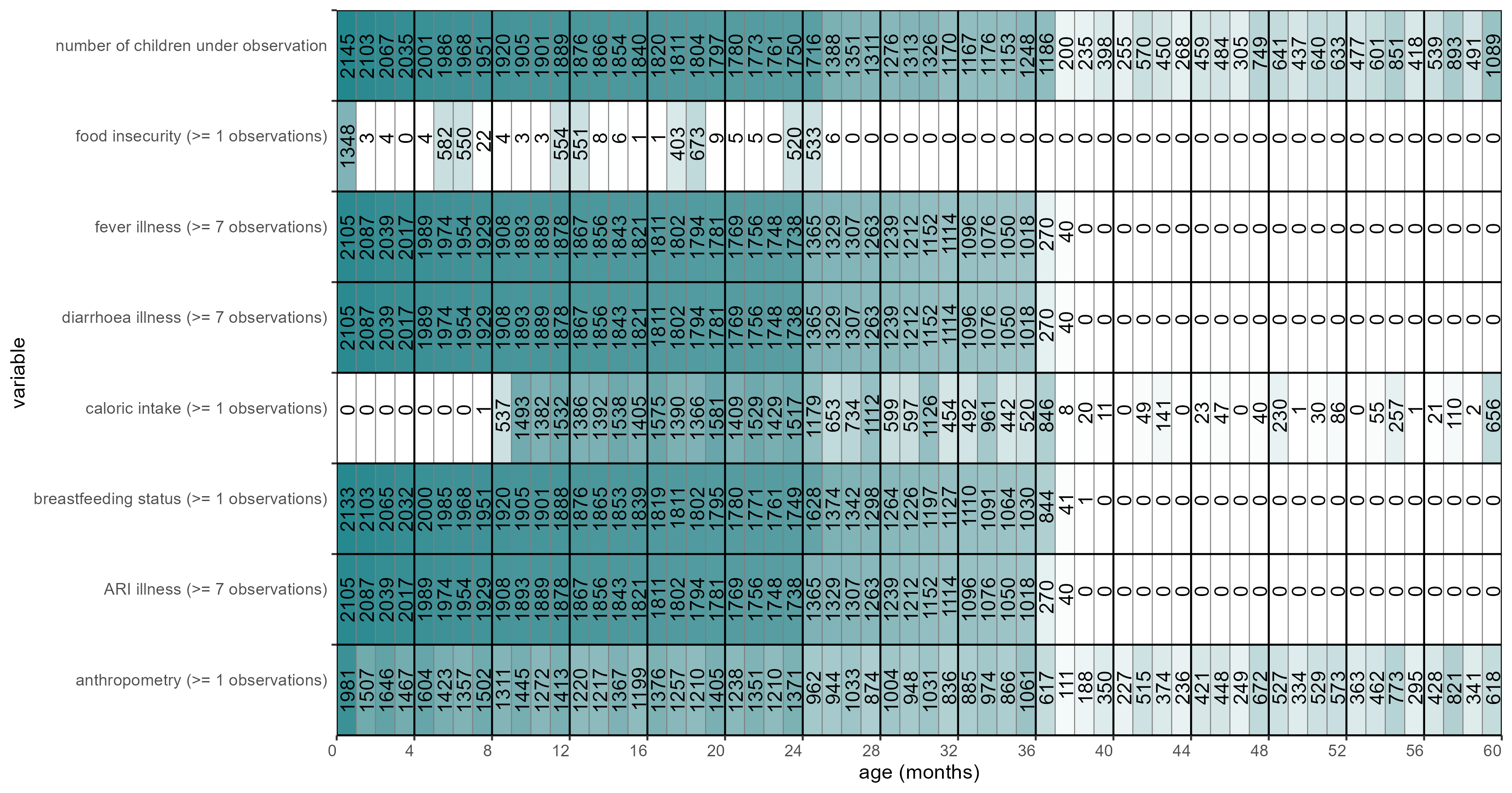

Figure S2. Availability of data, by variable and month in children’s life. Figures inside each cell indicate the number of available child observations with frequency fulfilling the analysis inclusion criteria (see specifications in parentheses after each variable name).

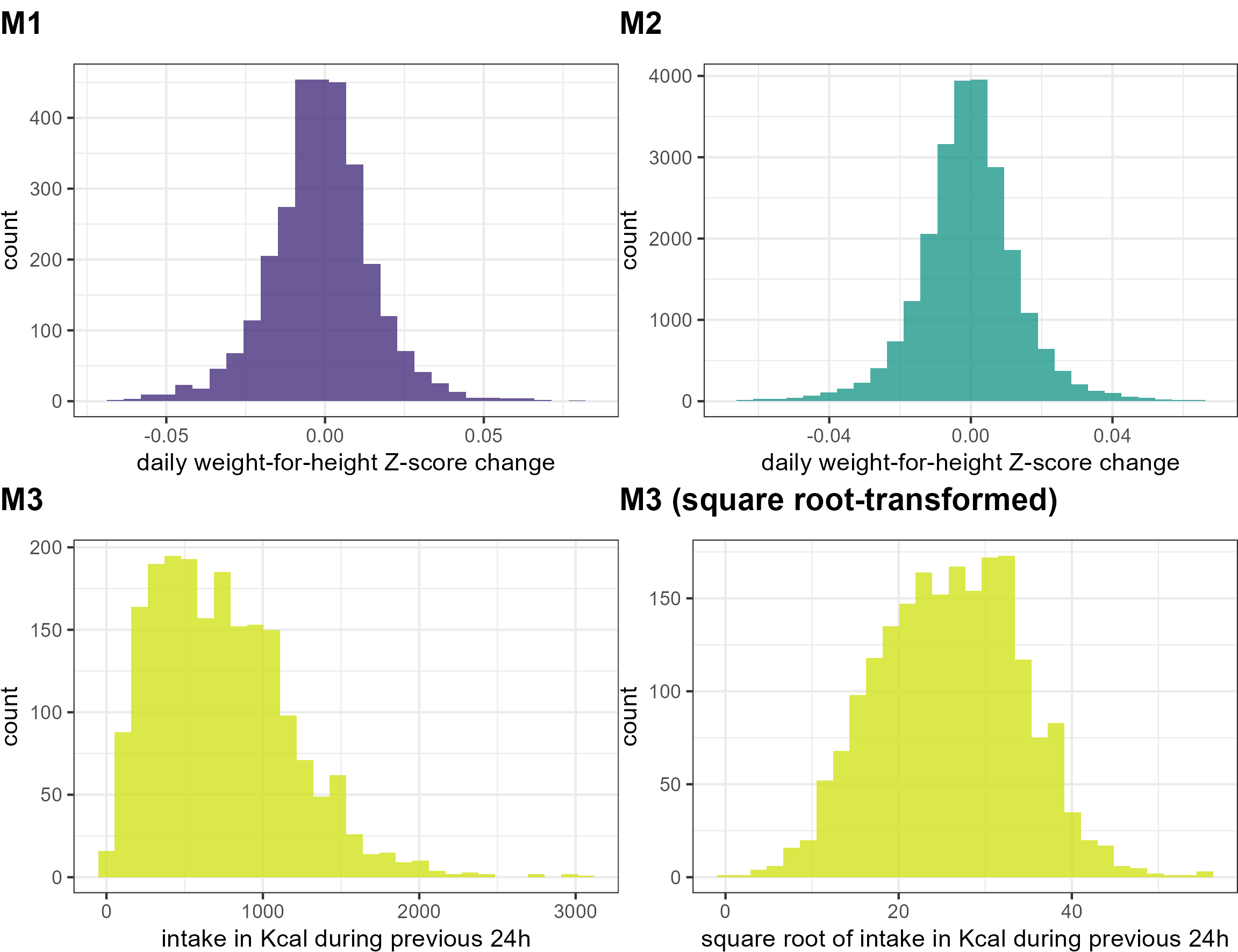

Figure S3. Distributions of observed values of the model outcomes.

### Model 1 evaluation results

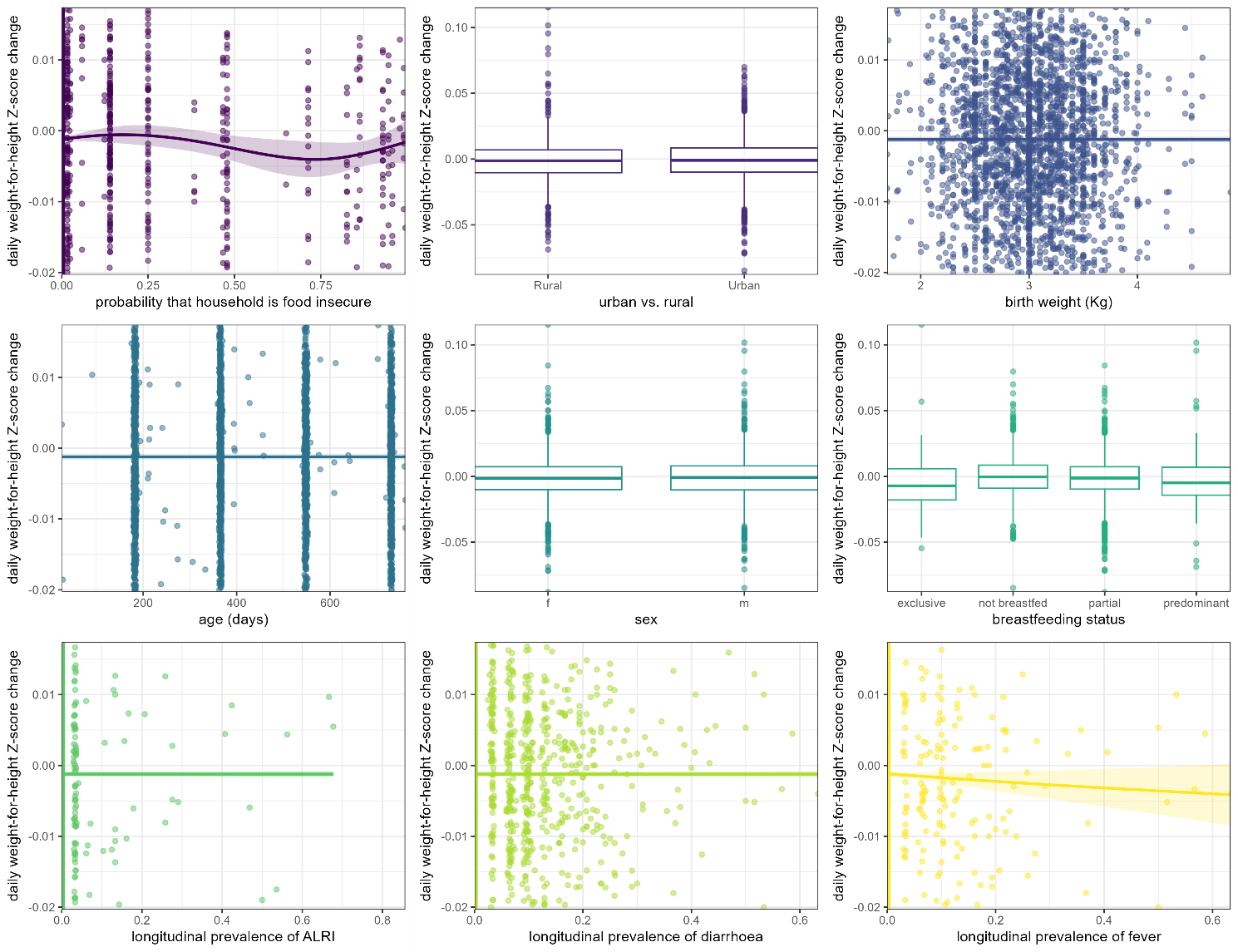

Figure S4. Correlation between each predictor and the outcome (M1). Lines and shaded areas indicate the point estimate and 95% confidence intervals of a generalised additive model smooth. Box plots for categorical predictors show the median, inter-quartile range (edges of box), 95% percentile interval (whiskers) and outliers (dots).

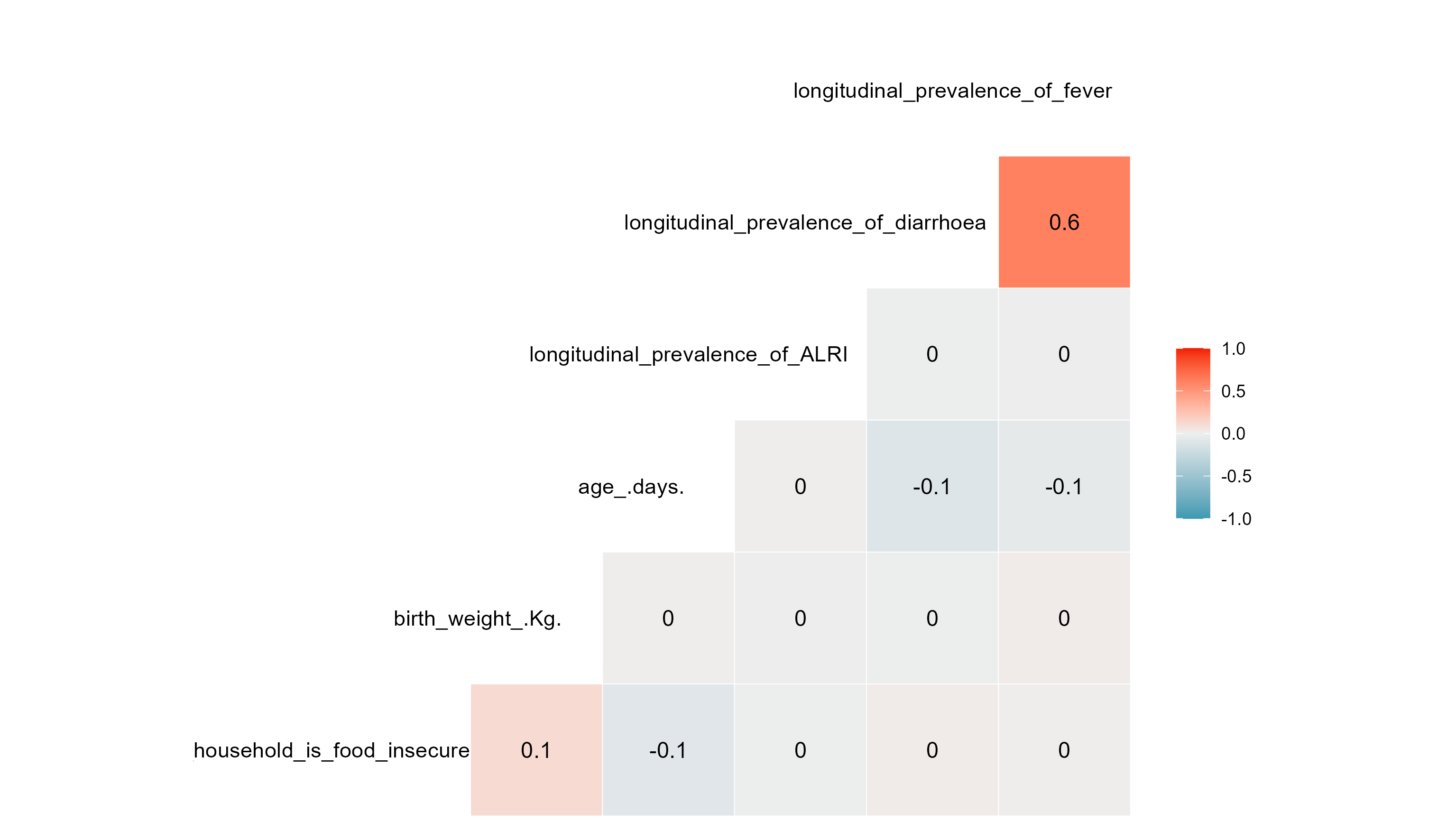

Figure S5. Pearson correlation coefficients for pairs of continuous predictors entered into M1.

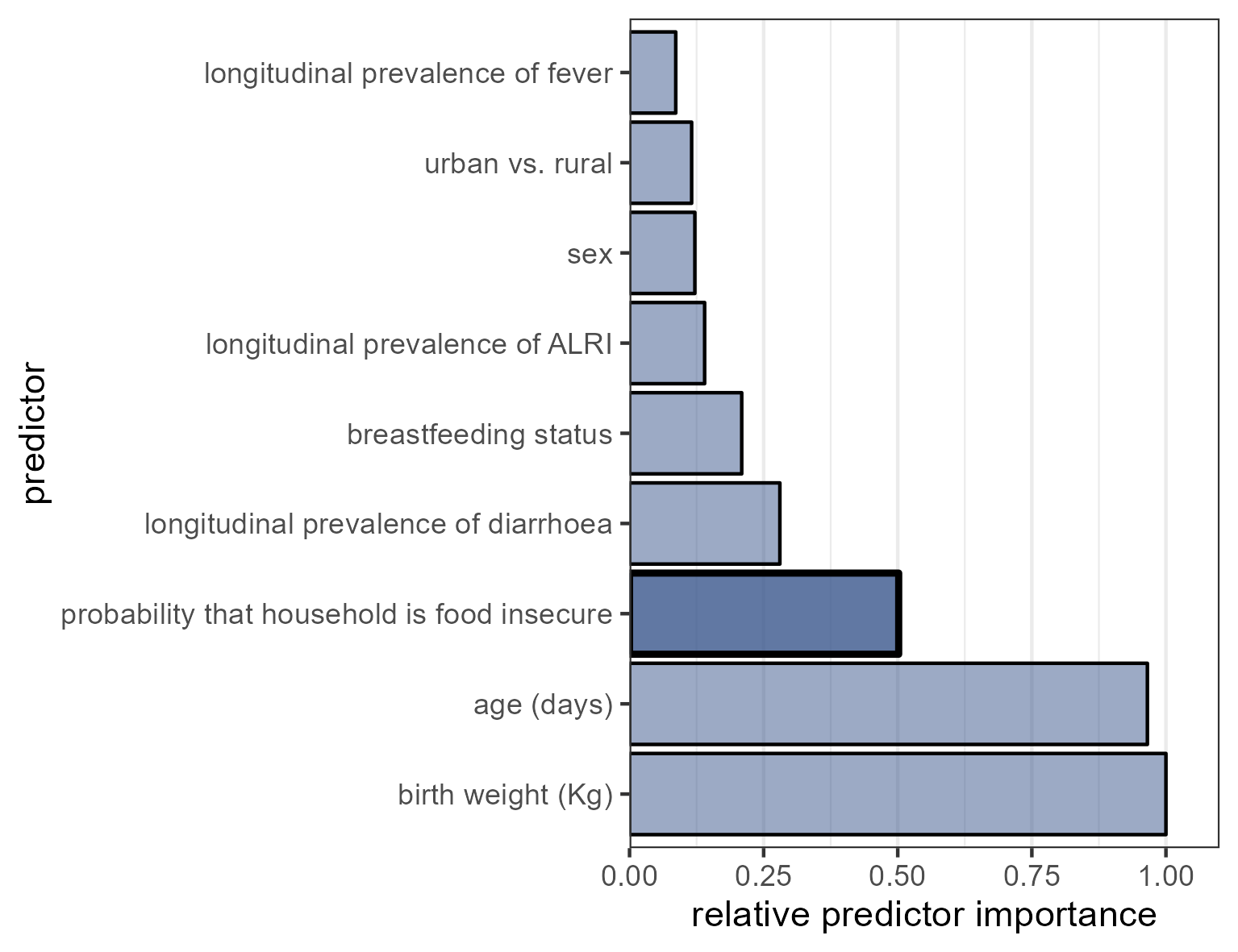

Figure S6. Relative importance of M1 predictors entered into a random forest algorithm.

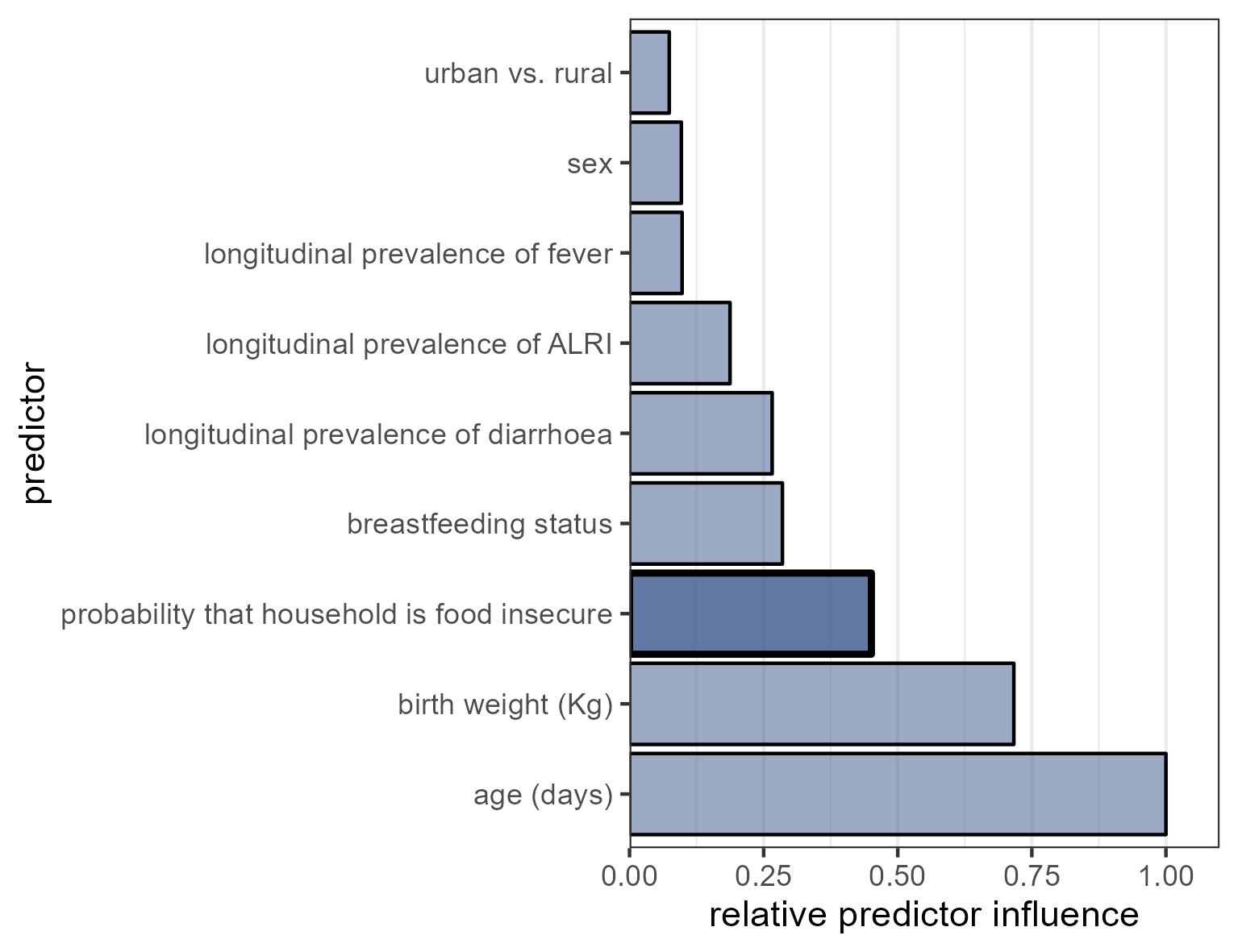

Figure S7. Relative influence of M1 predictors entered into a generalised boosted regression.

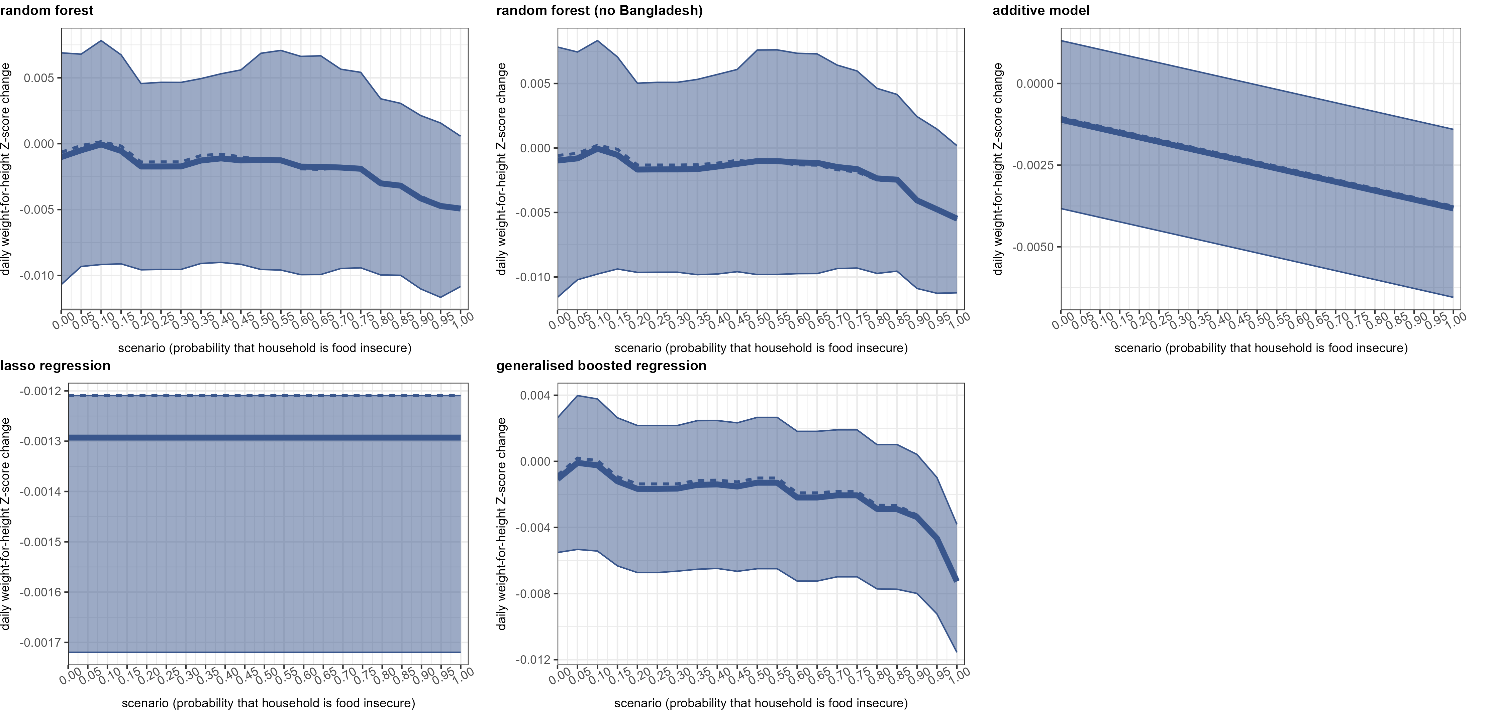

Figure S8. Predicted levels of the outcome under different hypothetical values of the main predictor of interest (as a multiplier of the original value in the data), by prediction method, for M1. The thick solid and dotted lines denote the mean and median predictions, with the shaded band encompassing the 95% confidence interval of the predictions.

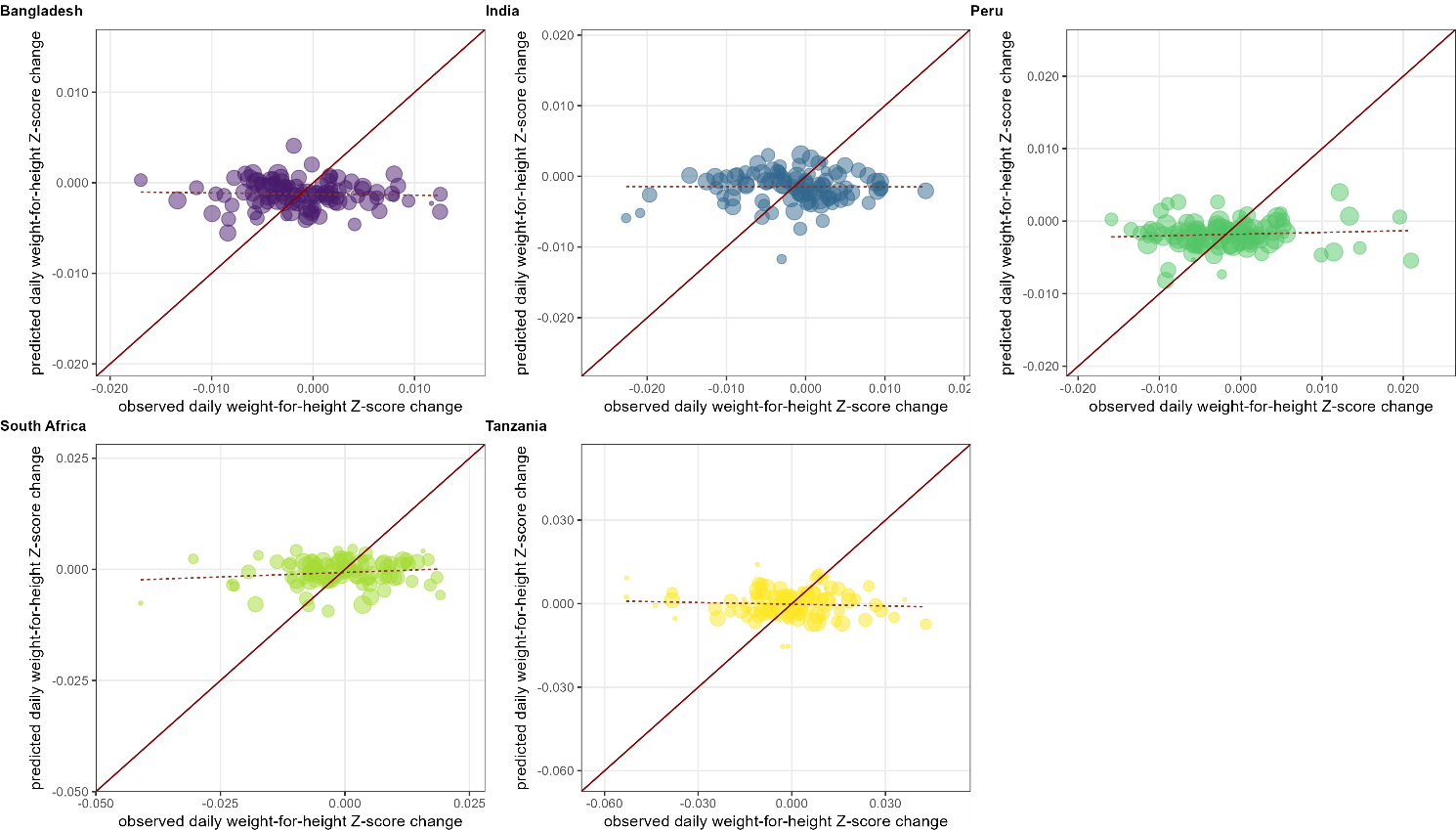

Figure S9. Mean random forest predictions versus observations, by cross-validation fold, for M1, when fitting a model for each country separately. Each dot represents a cross-validation fold, sized according to its relative share of the total dataset. The diagonal line indicates optimal prediction, while the dotted line indicates the slope of the predictions as a function of observations, based on a least-squares regression weighted by fold size.

### Model 2 evaluation results

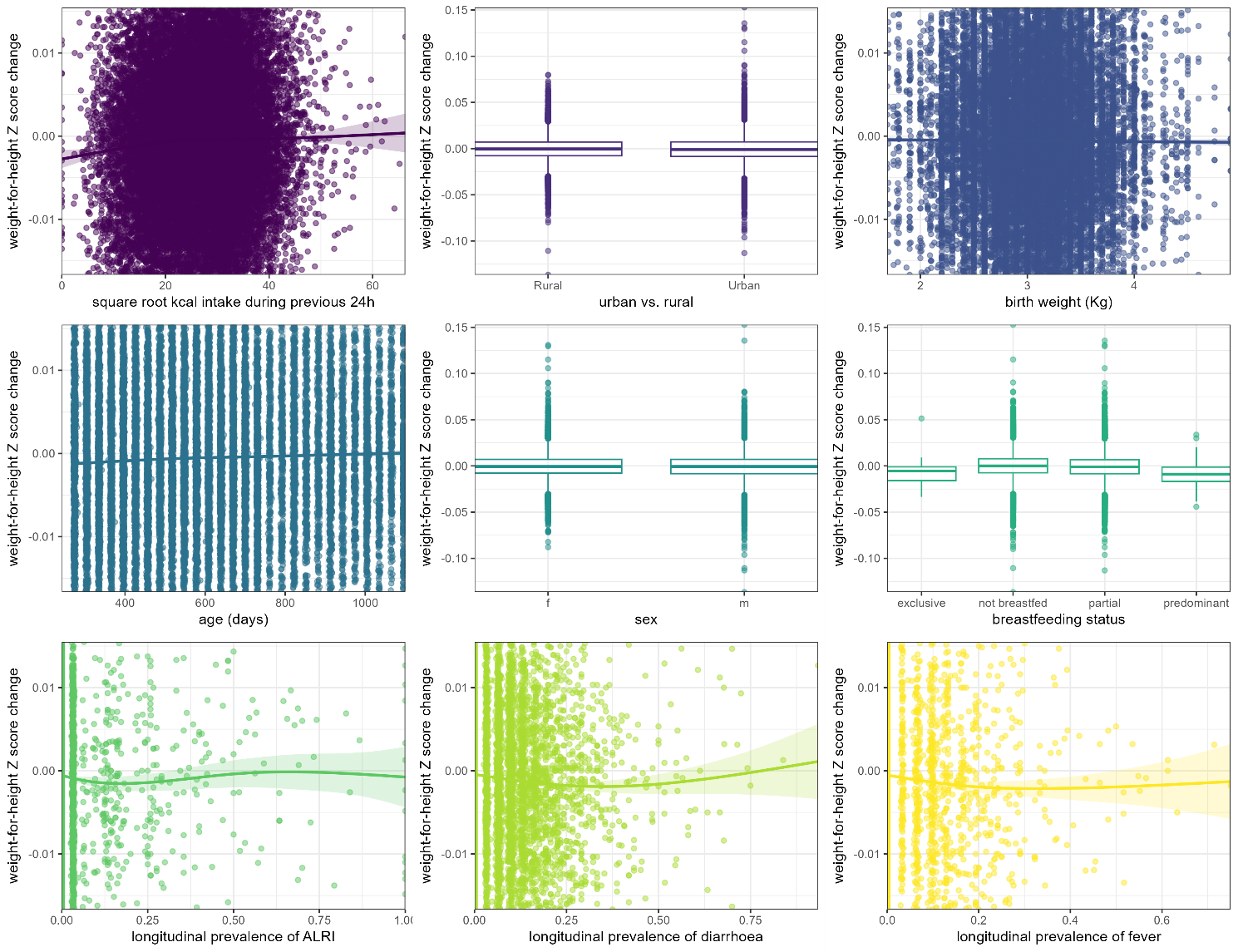

Figure S10. Correlation between each predictor and the outcome (Model 2). Lines and shaded areas indicate the point estimate and 95% confidence intervals of a generalised additive model smooth. Box plots for categorical predictors show the median, inter-quartile range (edges of box), 95% percentile interval (whiskers) and outliers (dots).

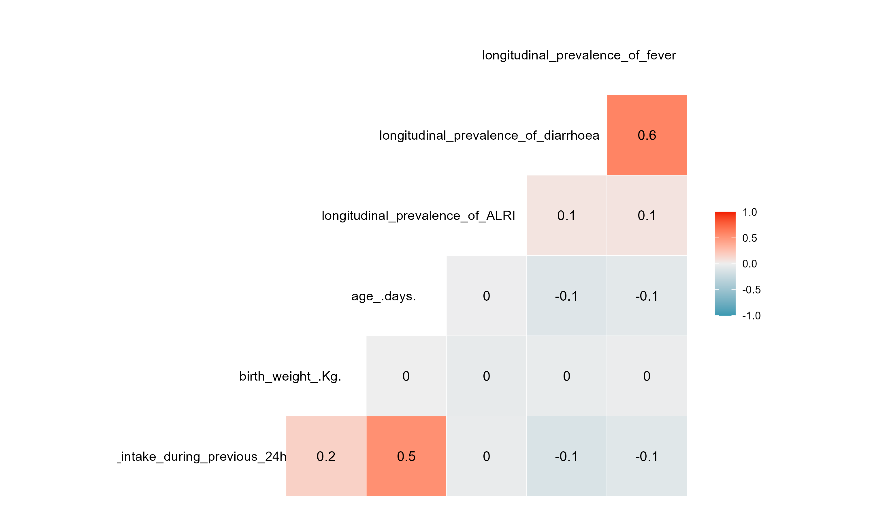

Figure S11. Pearson correlation coefficients for pairs of continuous predictors entered into M2.

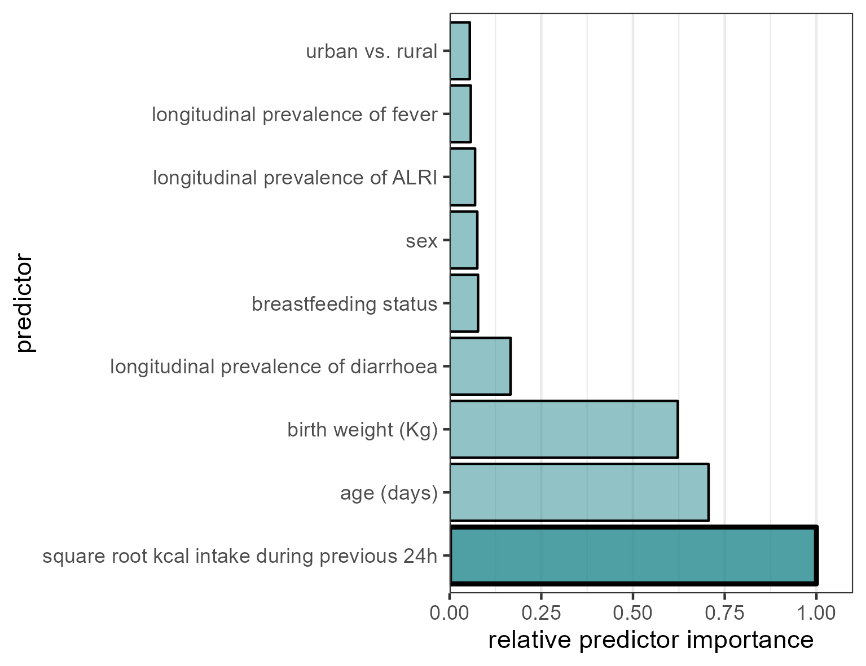

Figure S12. Relative importance of M2 predictors entered into a random forest algorithm.

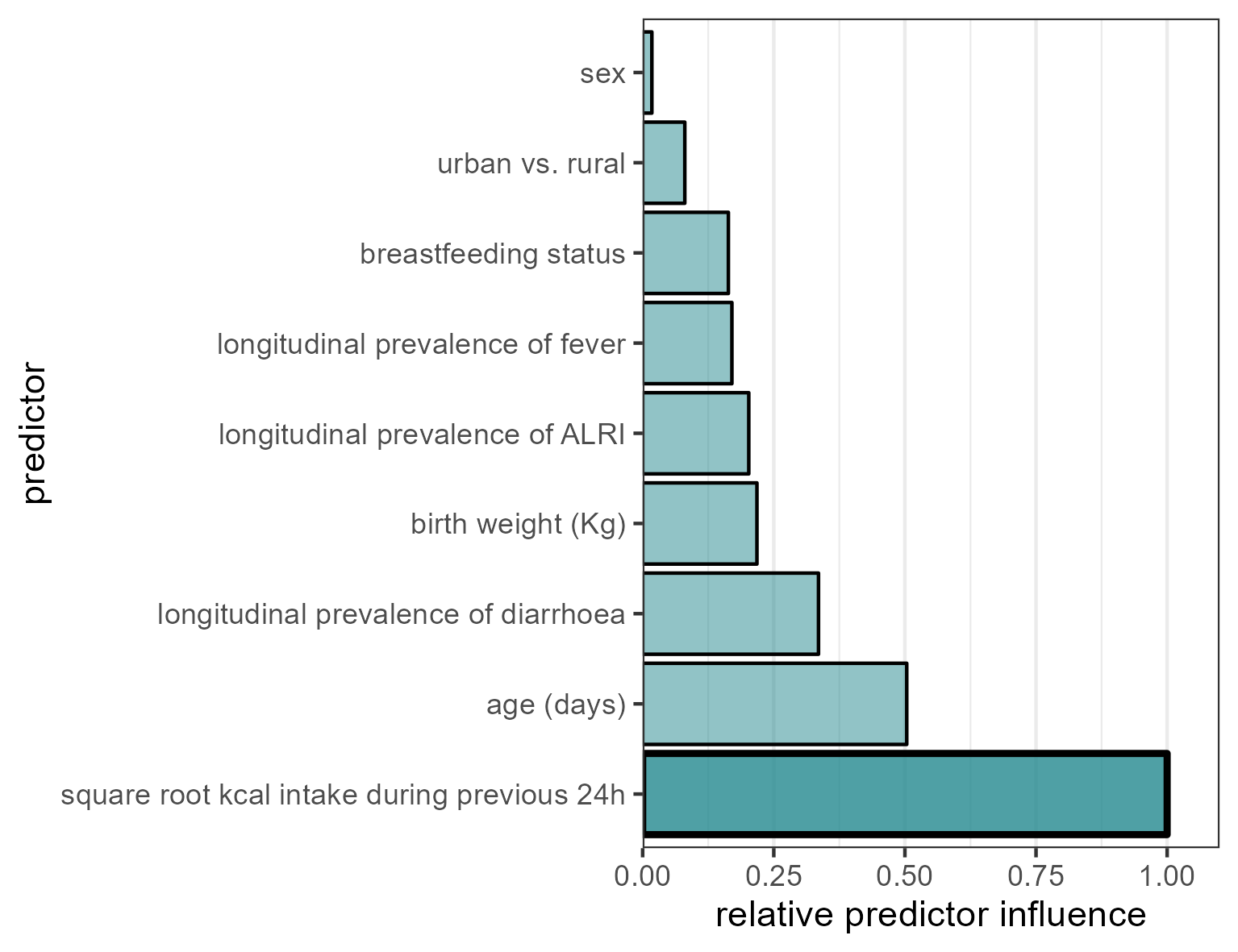

Figure S13. Relative influence of M2 predictors entered into a generalised boosted regression.

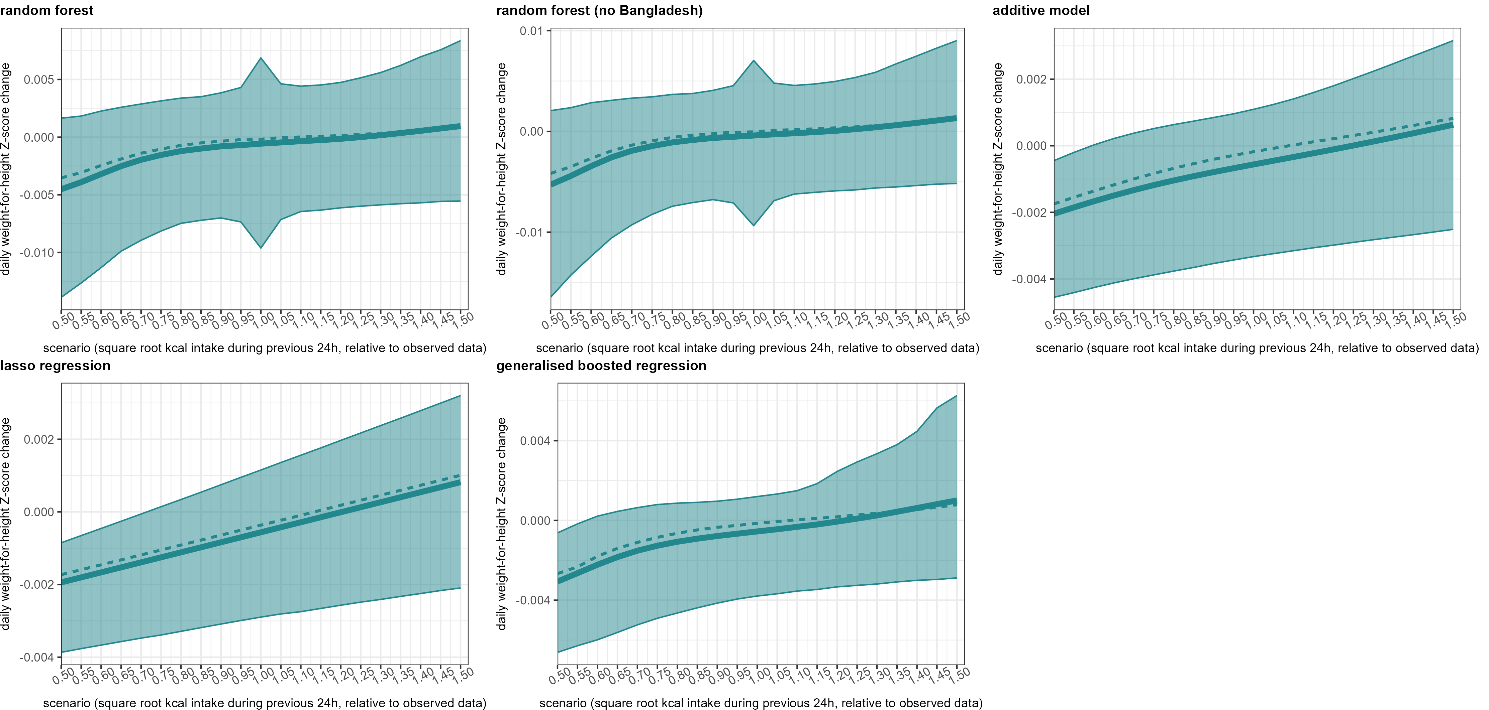

Figure S14. Predicted levels of the outcome under different hypothetical values of the main predictor of interest (as a multiplier of the original value in the data), by prediction method, for M2. The thick solid and dotted lines denote the mean and median predictions, with the shaded band encompassing the 95% confidence interval of the predictions.

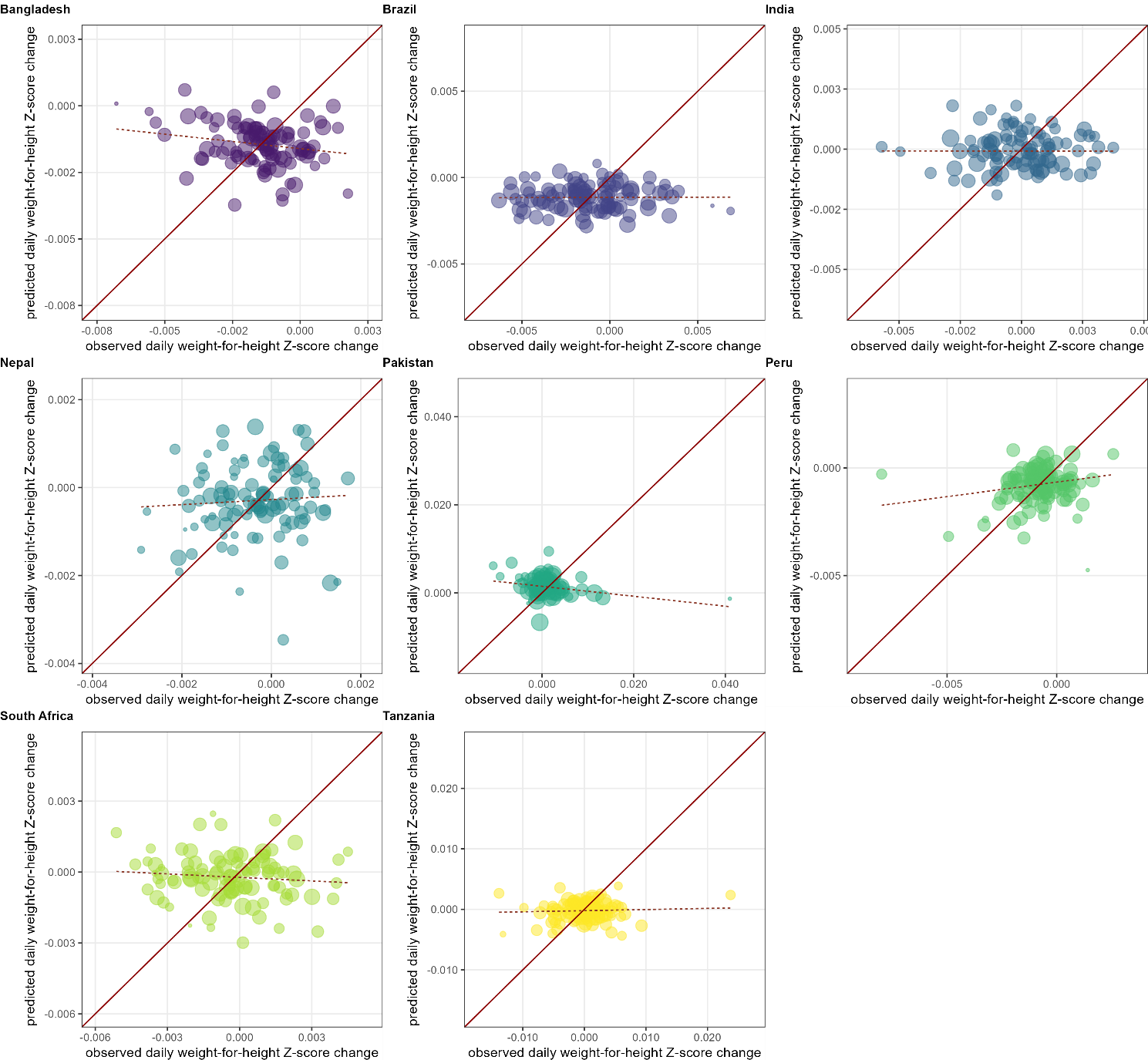

Figure S15. Mean random forest predictions versus observations, by cross-validation fold, for M2, when fitting a model for each country separately. Each dot represents a cross-validation fold, sized according to its relative share of the total dataset. The diagonal line indicates optimal prediction, while the dotted line indicates the slope of the predictions as a function of observations, based on a least-squares regression weighted by fold size.

### Model 3 evaluation results

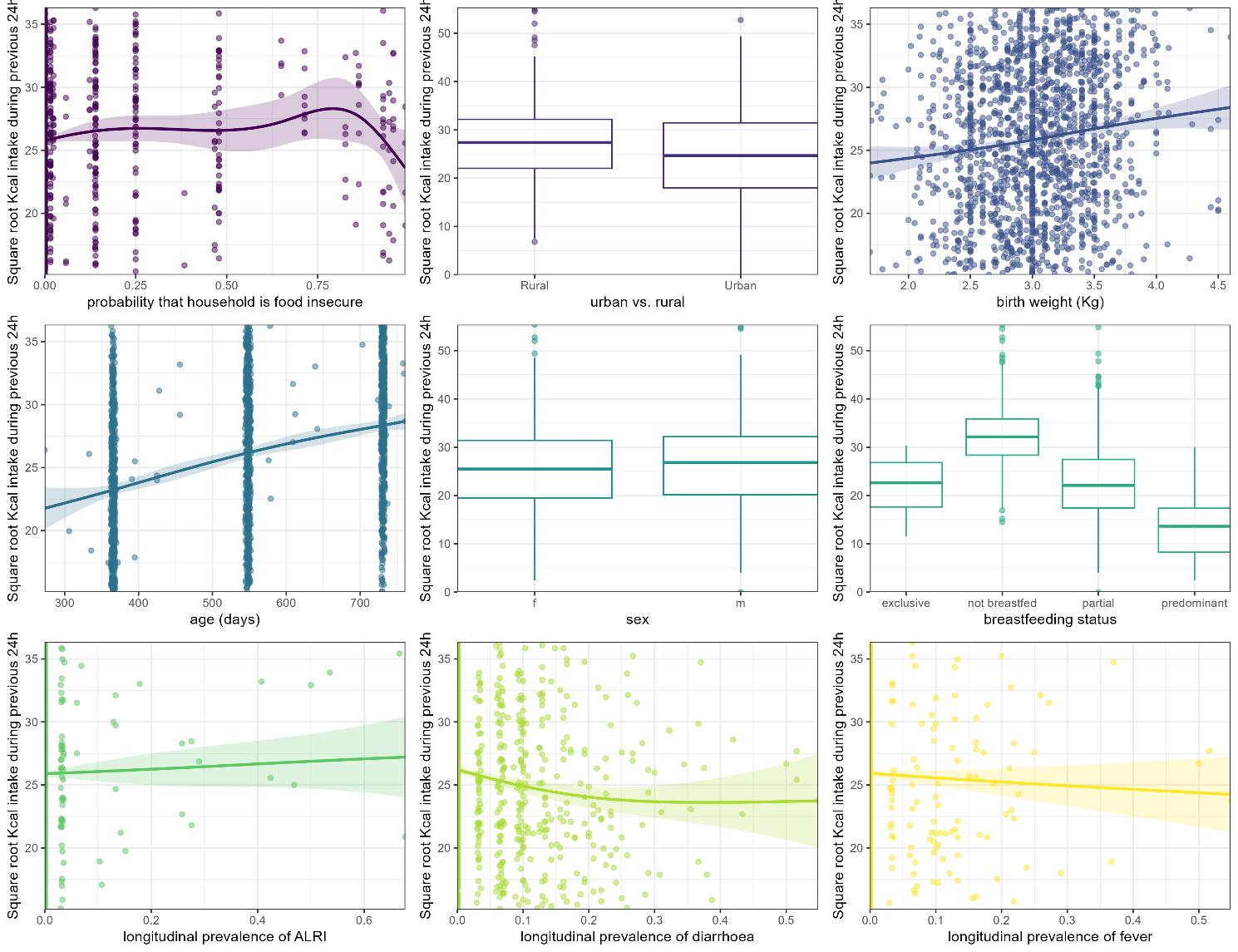

Figure S16. Correlation between each predictor and the outcome (Model 3). Lines and shaded areas indicate the point estimate and 95% confidence intervals of a generalised additive model smooth. Box plots for categorical predictors show the median, inter-quartile range (edges of box), 95% percentile interval (whiskers) and outliers (dots).

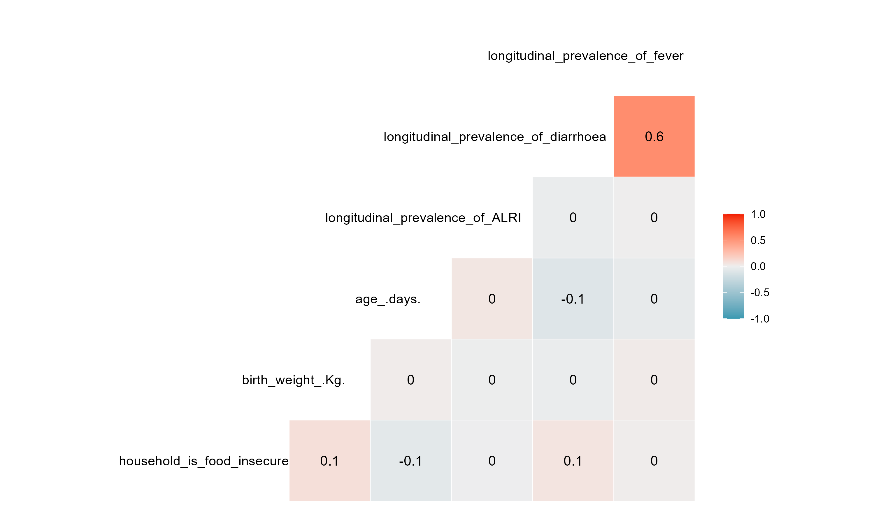

Figure S17. Pearson correlation coefficients for pairs of continuous predictors entered into M3.

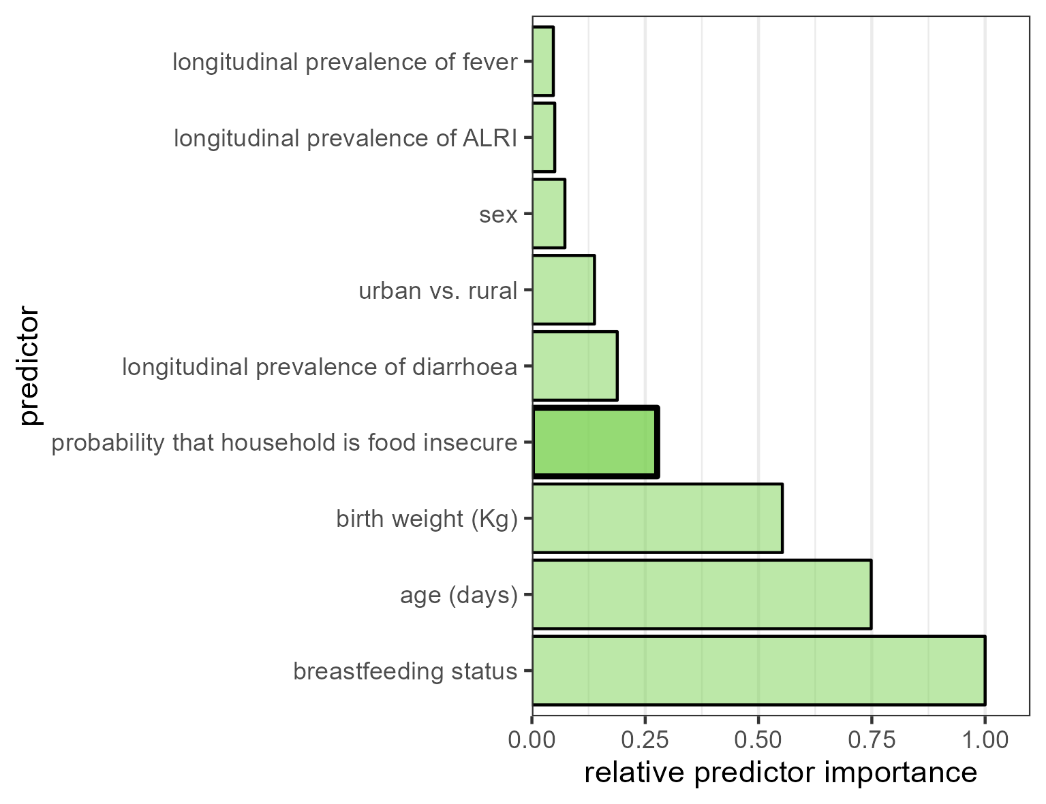

Figure S18. Relative importance of M3 predictors entered into a random forest algorithm.

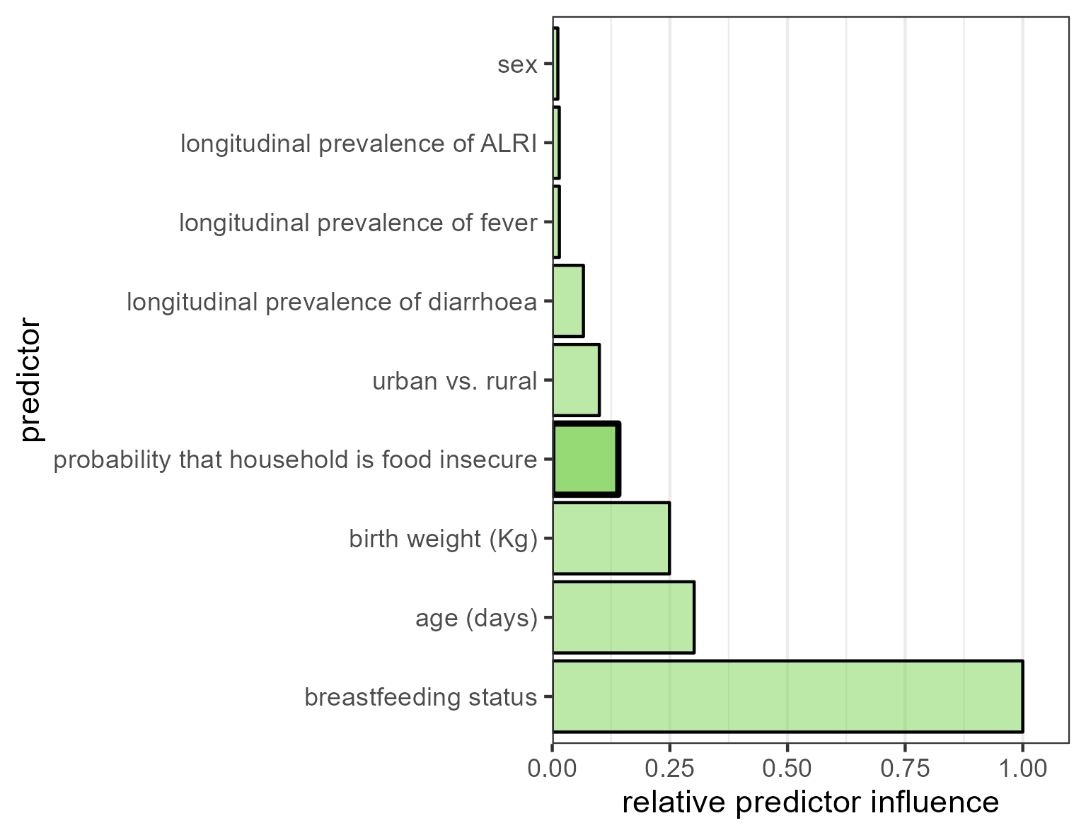

Figure S19. Relative influence of M3 predictors entered into a generalised boosted regression.

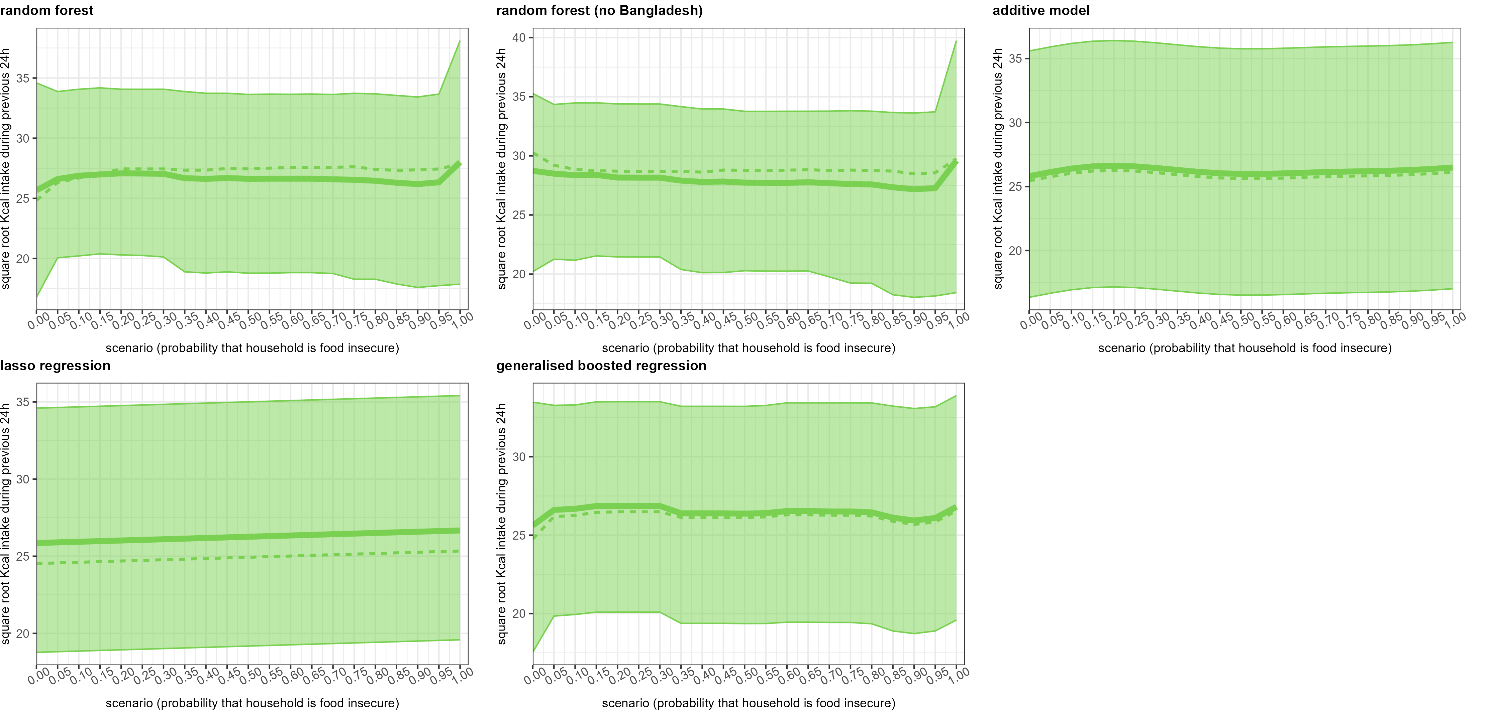

Figure S20. Predicted levels of the outcome under different hypothetical values of the main predictor of interest (as a multiplier of the original value in the data), by prediction method, for M3. The thick solid and dotted lines denote the mean and median predictions, with the shaded band encompassing the 95% confidence interval of the predictions.

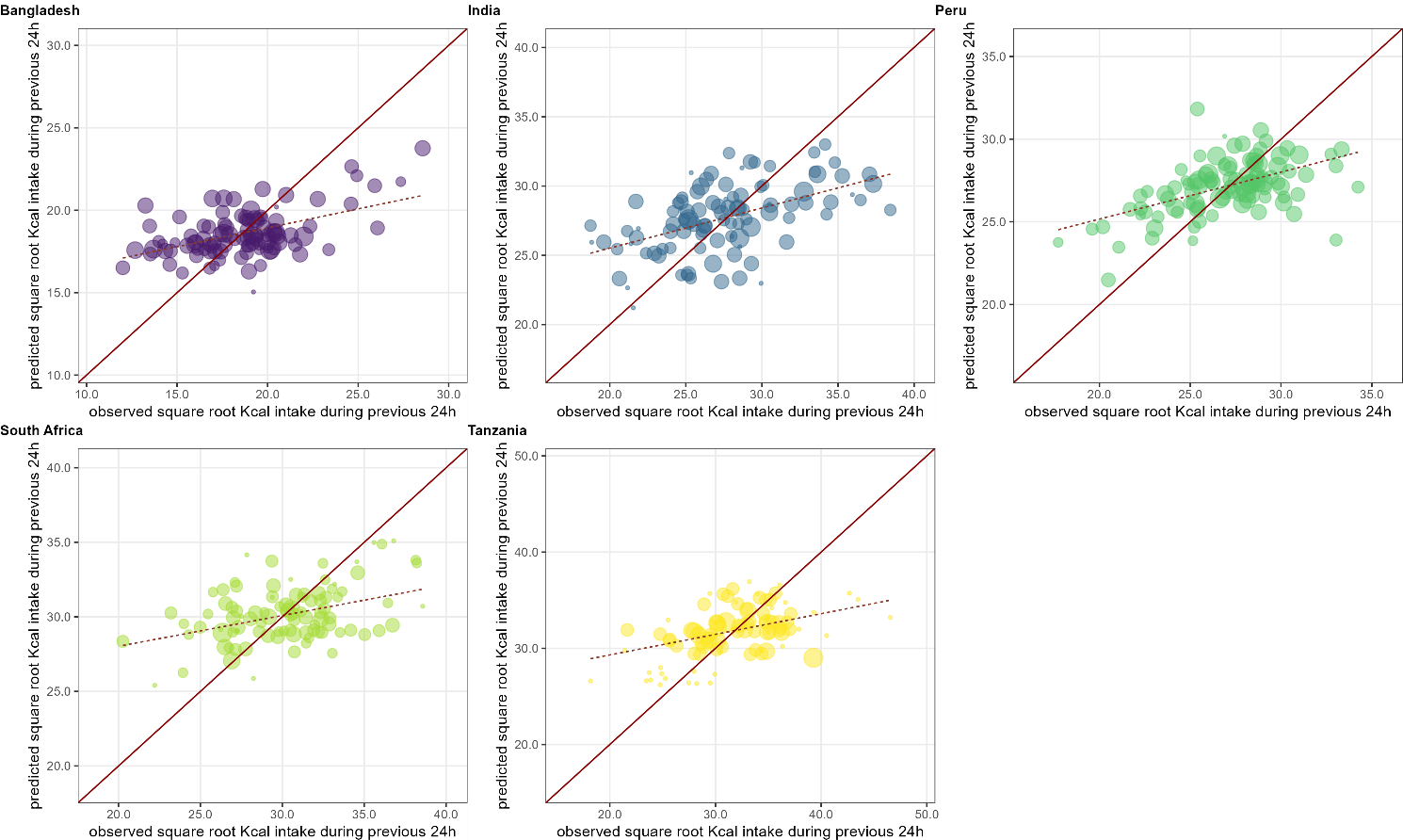

Figure S21. Mean random forest predictions versus observations, by cross-validation fold, for M3, when fitting a model for each country separately. Each dot represents a cross-validation fold, sized according to its relative share of the total dataset. The diagonal line indicates optimal prediction, while the dotted line indicates the slope of the predictions as a function of observations, based on a least-squares regression weighted by fold size.
